# Analyzing concordance between MUAC, MUACZ, and WHZ in diagnosing acute malnutrition among children under 5 in Somalia

**DOI:** 10.1101/2025.02.19.25322555

**Authors:** Sydney Garretson, Shelley Walton, Kemish Kenneth Alier, Samantha Grounds, Qundeel Khattak, Said Aden Mohamoud, Abdullahi Arays, Farhan Mohamed Mohamud, Abdullahi Muse Mohamoud, Adan Mahdi, Meftuh Omer, Mohamed Billow Mahat, Fabrizio Loddo, Marina Tripaldi, Nadia Akseer

**Author notes:** **Corresponding Author:** Nadia Akseer, Associate Scientist, Department of International Health Johns Hopkins University.

## Abstract

**Background:** Acute malnutrition, also known as wasting, affected an estimated 45 million children under 5 (CU5) globally in 2023. Wasting is measured using a child’s mid upper arm circumference (MUAC), weight-for-height z-score (WHZ), or nutritional edema. In low-resource contexts, MUAC is often the only measurement used to regularly screen for malnutrition, but recent research suggests MUAC alone fails to diagnose 25%-80% of WHZ-wasted children. MUACZ, an age-adjusted MUAC z-score, may identify additional cases missed by MUAC alone.

**Methods:** This was a secondary analysis of 1,408 CU5 enrolled in a cluster-randomized controlled trial in two high-wasting regions of Somalia. This analysis explored wasting prevalences, concordance between indicator pairs, linear regression modelling, and ROC analysis using WHZ as a gold standard to examine alternative MUAC and MUACZ thresholds.

**Results:** Wasting prevalence was 1.5% by MUAC, 8.5% by MUACZ, and 14.8% by WHZ. MUAC alone failed to identify 94% of WHZ-wasted children. There was slight concordance between MUAC and WHZ (κ=0.089) and fair concordance between MUACZ and WHZ (κ=0.385). Linear regression indicated that MUAC, age, sex, and stunting were all statistically significant variables for estimating child WHZ. Using WHZ as a gold standard, a MUAC of 13.7cm and MUACZ of –0.9 SD both accurately diagnosed >82% of wasted children ages 9-23 months. A MUAC of 14.4cm and MUACZ of −1 SD both accurately diagnosed >76% of wasted children ages 24-59 months.

**Conclusion:** Poor concordance between MUAC and WHZ should prompt review of wasting measurement guidelines for this population. Stratified analyses and regression modelling showed increased MUAC with age among CU5. MUACZ or age-specific MUAC thresholds may identify more truly wasted children, improving coverage of treatment interventions. Policymakers should examine how adapting screening guidelines impacts health facilities and treatment availability. Future studies should consider long-term outcomes and mortality associated with increased thresholds of MUAC and MUACZ.

**Registration:** The cluster-RCT is registered at ClinicalTrials.gov, ID: NCT06642012.

## 1. Background

### 1.1. Acute Malnutrition in Somalia

A recent UNICEF report includes Somalia among the 15 countries worldwide with the highest rates of malnutrition, food insecurity, and food poverty.[1] This humanitarian crisis in Somalia has seen increasing rates of wasting recently: the Integrated Food Security Phase Classification (IPC) estimates that in 2024, 1.7 million children under 5 (CU5) in Somalia will be acutely malnourished, 430,000 of whom will be severely wasted.[2]

### 1.2. Anthropometric Origins for Diagnosing Acute Malnutrition

The World Health Organization (WHO) has established guidelines for diagnosing acute malnutrition based on the presence of edema and two anthropometric criteria, mid-upper arm circumference (MUAC) and weight-for-height z-score (WHZ).[3] WHZ compares a child’s weight to another child of the same height from a standard reference population and is scored as the number of standard deviations from the median reference WHZ.[4] Non-malnourished CU5 have a MUAC ≥ 12.5cm <u>and</u> WHZ ≥ −2 SD. Moderate acute malnutrition (MAM) is defined as having 11.5 cm ≤ MUAC < 12.5cm and/<u>or</u> −3≤ WHZ< −2 SD. Severe acute malnutrition (SAM) is defined as having MUAC < 11.5cm <u>and/or</u> WHZ < −3 SD <u>and/or</u> the presence of nutritional bilateral edema. In accordance with Somalia’s 2021 Integrated Management of Acute Malnutrition (IMAM) Guidelines, community screening for malnutrition utilizes MUAC and/or edema as criteria for healthcare facility treatment referrals.[5] The child’s weight and height are measured upon admission to a health center for wasting treatment and during follow-up visits, but are not used for screening due to resource constraints, measurement complications, and limited awareness and policy enforcement from both government and non-government organizations.

MUAC was initially recommended in the 1960s for assessing malnutrition in CU5 due to its strong correlation with muscle mass, offering advantages over weight-for-height.[6–8] Over time, research revealed age-related variations in MUAC, prompting a shift towards weight-for-height as a more precise measure of child nutrition status.[6,9] However, the use of MUAC tape for malnutrition diagnosis persists in many low-resource settings and community health screenings due to its low cost and ease of measurement.[10,11] The threshold of MUAC <11.5cm for SAM diagnosis remains pivotal and is linked to increased mortality risk among CU5.[6,12,13]

Recent population-level representative surveys conducted in humanitarian settings have shown that MUAC wasting prevalences most often underestimate WHZ wasting prevalences by 5-10 percentage points.[14–17] In 2018, Bilukha and Leidman analyzed population-level survey data in humanitarian settings from 41 countries worldwide and found that 74% of surveys had a higher wasting prevalence by WHZ than by MUAC.[15] The greatest differences in wasting prevalences by indicator were found in regions with the highest rates of wasting by WHZ.[15]

MUAC and WHZ also diagnose different children as wasted. Leidman 2019 and Roberfroid 2015 found that among children identified as wasted by either MUAC or WHZ, only 15-30% were identified by both criteria.[14,18] Female, younger, and stunted children are more likely to be diagnosed as malnourished by MUAC.[18–21] Cohen’s kappa statistic, which ranges from 0 (poor agreement) to 1 (perfect agreement) has been used in several studies to assess concordance between MUAC and WHZ: concordance results ranged from 0.21 to 0.35, indicating fair concordance at best.[16,19,22] Previous research has also used receiver-operating characteristic (ROC) analysis to explore how increasing clinical cutoffs may improve MUAC’s sensitivity compared to WHZ: MUAC cutoffs greater than 13.5cm have shown increased sensitivity for diagnosing wasted CU5.[16,19,20]

MUACZ, an age-adjusted MUAC z-score, may be a useful measure to assess CU5 malnutrition and resolve discrepancies between MUAC and WHZ. The recent development of MUACZ tape also simplifies measurement procedures.[23] Prior studies have found that MUACZ categorized a higher percentage of children as wasted than both MUAC and WHZ, but the difference was not substantial enough to warrant programmatic use.[14,22] Custodio et al. analyzed surveillance data in Somalia to evaluate MUACZ; the prevalence of acute malnutrition was more similar by WHZ and MUACZ (∼16%) compared to 8% by MUAC only.[24]

Research on measurement concordance in Somalia has found more similar wasting prevalences using MUACZ and WHZ, with MUAC again underestimating the true burden of child wasting.[14,15,24] Analyses of global survey datasets have noted Somalia as an outlier due to substantial regional differences in malnutrition prevalence, finding poor concordance and an 8-10 percentage point difference in wasting prevalences by MUAC and WHZ.[15,24,25] However, there is a need to supplement this evidence with the current study because, to our knowledge, no studies have generated recent wasting prevalences in Somalia, especially among regions with high burdens of acute malnutrition. Other concordance studies have used national survey datasets and present population-level reviews of data collected from 1986 to 2022. This analysis uses anthropometric data collected from CU5 enrolled in a cluster-randomized controlled trial to generate real-time wasting prevalence estimates within 6 months of data collection.

### 1.3. Study Aims and Objectives

In Somalia, MUAC is the only in-field screening criteria for wasting diagnosis and admission to wasting treatment, so understanding concordance between MUAC and WHZ in this population is critical for programmatic use.

The aim of this study is to examine concordance between MUAC, MUACZ, and WHZ prevalence estimates of acute malnutrition among children under 5 in two high wasting burden regions in Somalia, Bay and Hiran. Specific objectives were to 1) Generate acute malnutrition prevalences using each anthropometric indicator (MUAC, MUACZ, and WHZ), 2) Compare the concordance and agreement between indicators for assessing child wasting (defined as MUAC < 12.5cm, MUACZ <−2, WHZ<−2) in children ages 6-59 months in Somalia, stratified by two regions (Bay vs Hiran) and by child age (9-23 months vs 24-59 months), and 3) Examine sensitivities and specificities of alternative MUAC and MUACZ thresholds using WHZ as a gold standard.

## 2. Methods

### 2.1 Study Setting

This study analyzed data from a cluster-randomized controlled trial conducted in two high wasting burden regions of Somalia, Bay and Hiran. These regions are heavily impacted by conflict and climate instability, leading to high rates of food insecurity and displacement from conflict, droughts, and flooding.[26–28]

Data for the trial were collected from May-December 2023 for the Cash Plus for Nutrition program implemented by Save the Children in Somalia. This cluster-randomized controlled trial enrolled CU5 and their mothers in Bay and Hiran to investigate which combinations of cash assistance are most effective and cost-effective for reducing malnutrition. Inclusion criteria for study enrollment were registration in a Bureau for Humanitarian Assistance (BHA)-funded program, no prior wasting by MUAC within the last 12 months, and no wasting by MUAC at baseline data collection (May 2023). A detailed description of the trial design can be found in the study’s methodology paper.[29]

### 2.2 Study Design

This is a secondary analysis of endline data (collected December 2023) emerging from the cluster-randomized trial. Data were analyzed as a cohort of children.

### 2.3 Outcomes

A full table of outcome definitions can be found in the Table S17. Child MUAC was measured twice using standard MUAC tape, to the nearest 0.1cm. Child weight was measured twice to the nearest 0.01kg, using a UNICEF electronic stand-on floor scale. Child height or length was measured twice to the nearest 0.1cm, using an ICLEAR Kid Growth Wooden Height-Length Measuring Board.

### 2.4 Analysis Approach

The initial dataset included 1,475 children ages 9-62 months. This analysis was restricted to CU5, so children ages 60-62 months were excluded (n=21). Using STATA’s *zanthro* module, the 2007 WHO Growth Standards were imported to transform anthropometric data into weight-for-height, height-for-age, and MUAC z-scores.[30] The 2007 WHO Growth Charts and *zanthro* command allowed us to calculate MUACZ for each child, which was not available using the 2006 WHO Growth Charts and the accompanying *zscore06* module in STATA.[30–32] Extreme or biologically implausible WHZ (<−5 SD and >5 SD) or height-for age (HAZ) values (<−6 SD and >6 SD) were dropped from the dataset (n=46).[33] The final analytical dataset included 1,408 children.

Wasting prevalences and 95% confidence intervals were calculated for wasting by MUAC, MUACZ, WHZ, and the 2013 WHO wasting management guidelines. Wasting prevalences were categorized as binary variables (wasted vs. not wasted) for each indicator, and additionally stratified into normal, MAM, and SAM.

Pearson’s correlation coefficients were used to assess linear correlation between anthropometric indicators. Linear regression was performed using WHZ as the outcome and MUAC, MUACZ, sex, age, and stunting as covariates. Stunting (HAZ <−2 SD) was included as a covariate of importance, since stunted children are especially vulnerable to malnutrition and may confound linear relationships between indicators.[18] Cohen’s kappa statistic was produced for pairwise combinations of MUAC, MUACZ, and WHZ to evaluate agreement in wasting diagnosis. Concordance was assessed with dichotomous diagnoses of wasting and with stratification into SAM/MAM/Normal.

Single-point ROC analysis was conducted using WHZ as a “gold standard”, examining sensitivities and specificities of different MUAC and MUACZ cutoffs for diagnosing wasting by WHZ. Criteria for determining “optimal” thresholds were maximizing AUC and sensitivity while maintaining a false positive rate less than 35%, similarly to previous analyses.[34]

The analysis was also stratified by child age and region to evaluate dose-response relationships since wasting prevalences varied significantly (Tables S5A and S6A). Age categories 9-23 months and 24-59 months were compared since this cutoff was used consistently in prior research.[14,17,20] Additionally, age was further stratified into four categories (9-23 months, 24-35 months, 36-47 months, and 48-59 months) to examine changes in child growth by year.

Two-sided alpha was set to 0.05 *a priori* and a p-value < 0.05 indicated statistical significance. All analyses were performed using STATA v 18.0 (StataCorp, College Station, TX, USA).

## 3. Results

The following results are from the endline dataset for this research study. Results were consistent from midline to endline with minimal differences in concordance and ROC analysis results. Midline results are included in the supplementary materials (Annexes S7-S14).

### 3.1. Descriptive Statistics and Table 1

Descriptive statistics were performed for demographic and anthropometric data (Table 1). There was a significantly larger proportion of female children in Bay than in Hiran. Mean child age and the proportion of children ages 24-59 months was greater in Hiran than in Bay. Mean MUAC, MUACZ, and WHZ were all lower in Hiran than Bay. Stunting prevalence was three times greater in Bay than in Hiran. There were moderate positive correlations between MUAC and WHZ (ρ=0.55), but stronger correlations between both MUAC and MUACZ, (ρ=0.85), and MUACZ and WHZ (ρ=0.65) (Figures S2A-S2C).

**Table 1.** Sample Characteristics.

| Sample Characteristics (N=1,408) |  |  |  |  |
| --- | --- | --- | --- | --- |
|  | Total Sample | Hiran (n=794) | Bay (n=614) | p values |
| Demographic Characteristics |  |  |  |  |
| Child Sex, n (%) |  |  |  |  |
| Male | 699 (49.6%) | 415 (52.3%) | 284 (46.3%) | 0.025 |
| Female | 709 (50.4%) | 379 (47.7%) | 330 (53.7%) |  |
| Child Age, mean (SD) | 36.82 (13.79) | 38.79 (13.72) | 34.27 (13.47) | <0.001 |
| 9-23 months (n, %) | 270 (19.2%) | 131 (16.5%) | 139 (22.6%) | 0.004 |
| 23-59 months (n, %) | 1,138 (80.8%) | 663 (83.5%) | 475 (77.4%) |  |
| Child Age - 4 categories* |  |  |  |  |
| 9-23 months (n, %) | 270 (19.2%) | 131 (16.5%) | 139 (22.6%) | <0.001 |
| 24-35 months (n, %) | 404 (28.7%) | 195 (24.6%) | 209 (34.0%) |  |
| 36-47 months (n, %) | 330 (23.4%) | 197 (24.8%) | 133 (21.7%) |  |
| 48-59 months (n, %) | 404 (28.7%) | 271 (34.1%) | 133 (21.7%) |  |
| Anthropometric Data, mean (SD) |  |  |  |  |
| Child MUAC (cm) | 14.73 (1.09) | 14.58 (1.04) | 14.91 (1.26) | <0.001 |
| Weight (kg) | 12.94 (2.57) | 13.33 (2.54) | 12.43 (2.53) | <0.001 |
| Height (cm) | 93.78 (10.88) | 96.50 (10.46) | 90.26 (10.41) | <0.001 |
| Z-Scores, mean (SD) |  |  |  |  |
| WHZ | -0.84 (1.10) | -1.09 (1.07) | -0.53 (1.06) | <0.001 |
| HAZ | -0.36 (1.51) | 0.026 (1.40) | -0.85 (1.52) | <0.001 |
| MUACZ | -0.76 (0.90) | -0.95 (0.86) | -0.53 (0.87) | <0.001 |
| Stunting Prevalence, n (%) |  |  |  |  |
| Not Stunted | 1243 (88.3%) | 744 (93.7%) | 499 (81.3%) | <0.001 |
| Stunted | 165 (11.7%) | 50 (6.3%) | 115 (18.7%) |  |
| Wasting Prevalences by Indicator, n (%) [95% CI] |  |  |  |  |
| MUAC | 21 (1.5%)<br>[1.0%, 2.3%] | 14 (1.8%)<br>[1.1%, 3.0%] | 7 (1.1%)<br>[0.6%, 2.4%] | 0.17 |
| WHZ | 208 (14.8%)<br>[13.0%, 16.7%] | 161 (20.3%)<br>[17.6%, 23.2%] | 47 (7.7%)<br>[5.8%, 10.1%] | <0.001 |
| Edema | 4 (0.3%)<br>[0.1%, 0.8%] | 2 (0.3%)<br>[0.1%,1.0%] | 2 (0.3%)<br>[0.1%, 1.3%] | 0.60 |
| 2013 WHO Wasting Guidelines:<br>by MUAC or WHZ or Edema | 218 (15.5%)<br>[13.7%, 17.5%] | 166 (20.9%)<br>[18.2%, 23.9%] | 52 (8.5%)<br>[6.5%, 11.0%] | <0.001 |
| MUACZ | 120 (8.5%)<br>[7.2%, 10.1%] | 91 (11.5%)<br>[9.4%, 13.9%] | 29 (4.7%)<br>[3.3%, 6.7%] | <0.001 |
| Wasting by any of the 4 Measures: (MUAC, WHZ, Oedema, or MUACZ) | 256 (18.2%)<br>[16.3%, 20.3%] | 192 (24.2%)<br>[21.4%, 27.3%] | 64 (10.4%)<br>[8.2%, 13.1%] | <0.001 |
| **Pearson's Correlations between Indicator Pairs, $\rho$** |  |  |  |  |
| MUAC & WHZ | 0.5501 | 0.4761 | 0.6095 |  |
| MUACZ & WHZ | 0.6458 | 0.5848 | 0.6771 |  |
| MUAC & MUACZ | 0.8455 | 0.8444 | 0.8437 |  |

\*For additional analyses, child age was stratified into 4 categories (9-23 months, 24-35 months, 36-47 months, and 48-59 months) to evaluate changes in optimal wasting thresholds per year of child development.

Distributions of child characteristics can be found in Figures S1A and S1B. 21 children were diagnosed as wasted by MUAC (1.5% of the sample). 208 children were diagnosed as wasted by WHZ (14.8%). 4 children had edema (0.3%). 218 unique children (15.5%) were identified as wasted according to the 2013 WHO guidelines.

Analyzing characteristics of wasted children (Table S1C), there was no statistically significant difference in MUAC wasting prevalence by child sex: 1.7% in male children vs 1.3% in female children (p=0.49). Male children (mean WHZ= −0.93, 95% CI: −1.01, −0.85) had a significantly lower mean WHZ than female children (mean WHZ = −0.76, 95% CI: −0.84, −0.68; p=0.003). Male children also had a significantly higher wasting prevalence by WHZ (16.9%, 95% CI: 14.10-19.65%) than female children (12.7%, 95% CI: 10.24-15.14%; p=0.03). Analyzing wasting by age group (Table S1D), 4.1% (95% CI: 1.72-6.43%) of younger children, ages 9-23 months, were wasted by MUAC compared to 0.9% of older children, ages 24-59 months (95% CI: 0.34-1.42%; p<0.001). Older children had on average lower WHZ (−0.89) than younger children (−0.65; p<0.001), but there was no statistically significant difference in wasting prevalence by WHZ between younger children (12.2%) and older children (15.4%; p=0.12). The total wasting prevalence using the 2013 WHO guidelines was significantly different between regions (p<0.001): 8.5% in Bay (95% CI: 6.27-10.67%; 52 out of 562 children) and 20.9% in Hiran (95% CI: 18.08-23.74%; 166 out of 628 children) (Table S5A).

When using MUACZ as an alternative wasting indicator, 120 children were diagnosed as wasted by MUACZ alone (8.5%). 256 unique children were identified as wasted by any of the 4 indicators (MUAC, WHZ, oedema, or MUACZ), yielding a prevalence of 18.2% (95% CI: 16.25-20.29%). Although the prevalence of wasting including MUACZ identified 38 additional children as wasted, this was not significantly different from the wasting prevalence by WHO guidelines (p=0.06).

### 3.2. Linear Regression Modelling

To explore relationships between WHZ and key anthropometric variables, linear regression was performed using WHZ as the outcome and MUAC, MUACZ, sex, age, stunting, and region as covariates. Table 2 presents regression modelling results with statistically significant covariates (Model 5). All other model results can be found in Table S3A (Models 1-7).

**Table 2.** Linear Regression Modelling Results – Model 5.

| **Model 5 ($R^2 = 0.46$)** |  |  |  |
| --- | --- | --- | --- |
|  | B (SE) | 95% Confidence Interval | p-value |
| MUAC | 0.66 (0.02) | (0.62, 0.77) | <0.001 |
| Age | -0.03 (0.01) | (-0.03, -0.02) | <0.001 |
| Sex <sup>a</sup> | 0.15 (0.04) | (0.07, 0.24) | <0.001 |
| Stunting <sup>b</sup> | 0.60 (0.07) | (0.47, 0.74) | <0.001 |
| Region <sup>c</sup> | 0.14 (0.05) | (0.04, 0.23) | 0.003 |

<sup>a</sup> 1= Female; <sup>b</sup> 1 = Stunted; <sup>c</sup> 1 = Bay.

In a simple linear regression of WHZ on continuous MUAC (Model 1), each centimeter change in MUAC was associated with a 0.56-unit increase in child WHZ (p<0.001, R^2^=0.30), and a MUAC of 12.5cm was associated with a WHZ of −2.08 SD. Simple linear regression of WHZ on additional covariates, age (continuous), sex, and stunting (yes/no), warranted their inclusion in a multiple linear regression as statistically significant covariates. Model 2 included both MUAC and age, showing age as the most significant confounder of the relationship between MUAC and WHZ, resulting in a 17.4% change in the regression coefficient. Model 3 included all covariates with age as a continuous variable, and after adjustment, a MUAC=12.5cm was associated with WHZ=−1.46. Model 4 demonstrated no evidence of interaction between age and sex in the adjusted model. In Model 5, when region (Bay vs Hiran) was added, the region coefficient was statistically significant (p=0.003) but did not confound relationship between WHZ and any other covariates.

Linear regression Models 6 and 7 examined MUACZ. In Model 6, a 1-unit change in MUACZ was associated with a 0.788-unit change in WHZ, and a MUACZ=−2 was associated with WHZ=−1.82 (p<0.001, R^2^=0.42). In the adjusted Model 7, age (p=0.89) and sex (p=0.12) were no longer statistically significant contributions, although stunting remained statistically significant (p<0.001).

### 3.3. Cohen’s Kappa and Concordance between Indicator Pairs

Concordance values for each pair can be found in Table S4A. MUAC and WHZ together identified 216 unique children as wasted: 195 by WHZ only, 8 by MUAC only, and 13 by both MUAC and WHZ (Table S4E). Treating WHZ as a ‘gold standard’ since it identifies more children as wasted, MUAC has a sensitivity of 6.25% and a specificity of 99.33%. Cohen’s kappa for agreement between MUAC and WHZ was 0.089, indicating only slight concordance between these two wasting indicators. When stratifying the wasted group into MAM and SAM, MUAC identified 0 children as SAM and 21 children as MAM; WHZ identified 24 children as SAM and 184 children as MAM (Table S4F).

MUAC and MUACZ together identified 121 unique children as wasted: 100 by MUACZ only, 1 by MUAC only, and 20 by both indicators Table S4G). Treating MUACZ as a gold standard, since it identifies more children as wasted, MUAC has a sensitivity of 16.67% and a specificity of 99.92%. Cohen’s kappa for agreement between MUAC and MUACZ is 0.265, indicating fair concordance between these two wasting indicators. After stratifying the wasting group into MAM and SAM, MUACZ identified 10 children as SAM and 110 children as MAM (Table S4H).

MUACZ and WHZ together identified 254 unique children as wasted: 134 by WHZ only, 46 by MUACZ only, and 74 by both indicators (Table S4I). Treating WHZ as a gold standard, MUACZ has a sensitivity of 35.58% and a specificity of 96.17%. Cohen’s kappa for agreement between MUACZ and WHZ was 0.385, indicating fair concordance between these two wasting indicators.

Stratifying the analysis by age group led to changes in concordance between each pair of wasting indicators (Table S4D). Among children 9-23 months, Cohen’s kappa for MUAC and WHZ = 0.225 (fair concordance), MUAC and MUACZ = 0.758 (substantial concordance), and MUACZ and WHZ = 0.278 (fair concordance). Among children 24-59 months, Cohen’s kappa for MUAC and WHZ = 0.060 (slight concordance), MUAC and MUACZ = 0.160 (fair concordance), and MUACZ and WHZ = 0.403 (moderate concordance).

### 3.4. Single-point ROC Analysis and Threshold Determination

Single-point ROC analysis allowed us to compare current MUAC and MUACZ thresholds to a more ideal threshold that would improve their sensitivity for diagnosing wasting, using WHZ as the gold standard. The goal of this analysis was to maximize the area under the curve (AUC) and sensitivity, since treating more children who are potentially wasted would be preferred over excluding truly wasted children. “Optimal” thresholds maximized AUC and sensitivity, while maintaining a false position rate less than 35% (specificity > 65%). Table 3 below presents the ROC analysis results.

**Table 3.** Full Sample Single-Point ROC Analysis Results using WHZ as a gold standard.

| **Full Sample (N=1,482)** |  |  |  |
| --- | --- | --- | --- |
| **MUAC** | **Sensitivity** | **Specificity** | **AUC** |
| MUAC < 12.5cm | 6.3% | 99.3% | 0.528 |
| MUAC < 14.4cm | 79.8% | 68.3% | 0.741 |
| **MUACZ** | **Sensitivity** | **Specificity** | **AUC** |
| MUACZ < -2 SD | 35.6% | 96.2% | 0.659 |
| MUACZ < -1 SD | 84.1% | 68.0% | 0.761 |

For the entire study sample, the current MUAC threshold of 12.5cm yielded sensitivity = 6.3%, specificity = 99.3%, and AUC = 0.528, indicating discrimination between cases and non-cases of WHZ wasting that is only slightly better than by chance alone. The optimal MUAC threshold for the sample was 14.4 cm, yielding a sensitivity = 79.8%, specificity = 68.3%, and AUC = 0.741. For a MUACZ threshold of −2, sensitivity = 35.6%, specificity = 96.2%, and AUC = 0.659. The optimal MUACZ threshold for the sample was −1, yielding sensitivity = 84.1%, specificity = 68.0%, and AUC = 0.761. Given the difference in wasting prevalence between the Bay and Hiran regions of the study, we stratified the analysis to examine potential differences in the recommended MUAC or MUACZ threshold by region, but there were not major differences in optimal cutoffs by region (Tables S5A & S5B).

We also conducted ROC analysis stratifying by age at 24 months (Tables S6A & S6B). Among children ages 9-23 months, the current MUAC threshold of 12.5 cm produces sensitivity = 18.2%, specificity = 97.9%, and AUC = 0.580. The optimal MUAC threshold for children ages 9-23 months was 13.7cm, yielding sensitivity = 81.8%, specificity = 73.4%, and AUC = 0.774. Among children ages 24-59 months, the current MUAC threshold of 12.5cm produces sensitivity = 4.0%, specificity = 99.7%, and AUC = 0.518. The optimal MUAC threshold for children ages 24-59 months was 14.4 cm, yielding sensitivity = 76.0%, specificity = 73.8%, and AUC = 0.749.

Among children ages 9-23 months, the current MUACZ threshold of −2 SD produced sensitivity = 18.2%, specificity = 97.9%, and AUC = 0.580. The optimal MUACZ threshold for children ages 9-23 months was −0.9, yielding sensitivity = 84.9%, specificity = 75.7%, and AUC = 0.802. Among children ages 24-59 months, the current MUACZ threshold of −2 produced sensitivity = 37.7%, specificity = 96.0%, and AUC = 0.668. The optimal MUACZ threshold for children ages 24-59 months was −1, yielding sensitivity = 85.1%, specificity = 65.3%, and AUC = 0.752.

ROC analysis was also stratified by age into four categories: 9-23 months, 24-35 months, 36-47 months, and 48-59 months. Optimal MUAC and MUACZ thresholds for each age category and performance results relative to WHZ are found in Table 4 below.

**Table 4.**
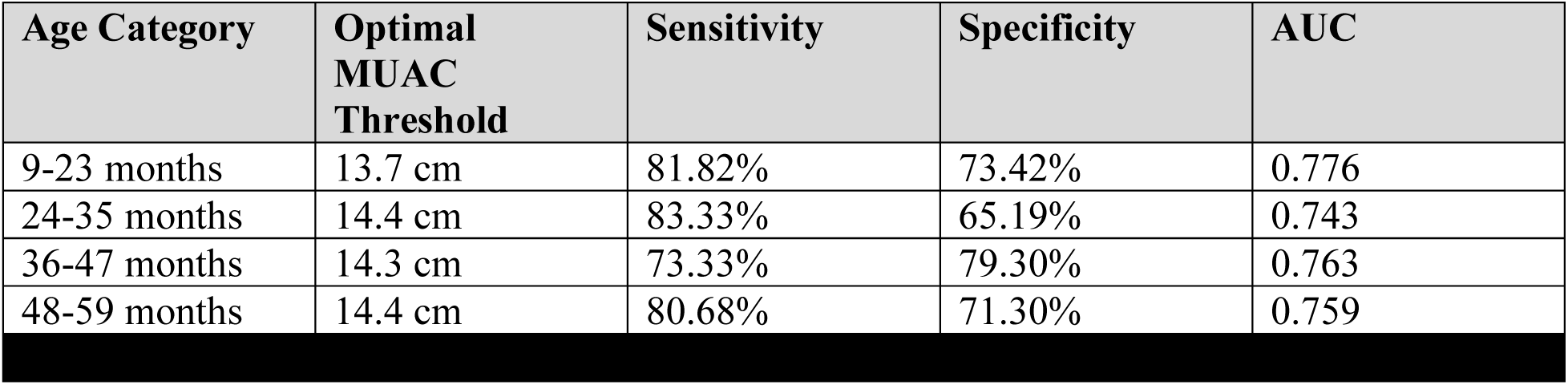
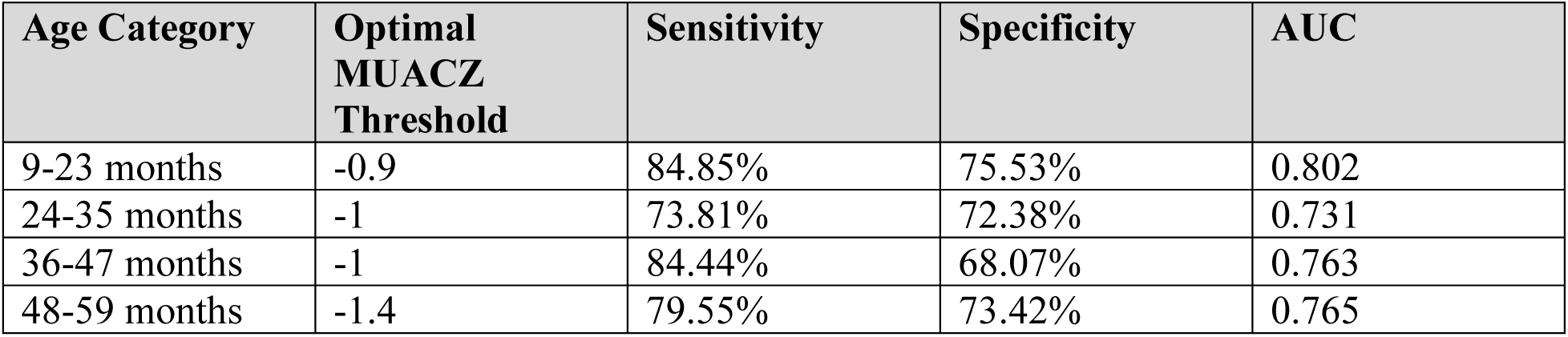
Optimal MUAC & MUACZ Thresholds, Sensitivity, Specificity.

## 4. Discussion

### 4.1 Summary

To our knowledge, this analysis is the first comparative assessment of MUAC, MUACZ, and WHZ diagnostic performance in two high wasting burden regions of Somalia. Our results indicate that the current MUAC threshold of 12.5cm to diagnose wasting heavily underestimates the number of children diagnosed as wasted by WHZ< –2 SD, yielding a prevalence difference of 13 percentage points. MUAC alone failed to diagnose 94% of the total WHZ-wasted children, missing 100% of children SAM by WHZ (n=24) and 94% of children MAM by WHZ (n=208). In addition, minimal concordance (κ=0.089) between current MUAC and WHZ cutoffs shows that these two indicators diagnose different children as wasted: male children were more likely to be diagnosed as wasted by WHZ, while younger children (ages 9-23 months) were more likely to be diagnosed as wasted by MUAC.

Twenty children (9.2% of the total wasted children using WHO guidelines) were identified as SAM by WHZ but normal by MUAC, and 175 children (80.3% of the total wasted children using WHO guidelines) were identified as MAM by WHZ but normal by MUAC. This discrepancy is alarming, since these children may not be referred for wasting treatment outside of this study due to reliance on MUAC alone for screening in Somalia. The 20 children who are severely malnourished are at extremely high risk for additional health complications or death, but their vulnerable status is not captured by their MUAC measurement alone. The current MUAC cutoff’s poor sensitivity (6.25%) for identifying children wasted by WHZ is insufficient to be used as a sole diagnostic indicator. Previous research has evaluated the use of MUACZ as an alternative indicator to measure wasting, with disagreement about its programmatic utility.[14,35] We evaluated adjusted MUAC thresholds and MUACZ to consider feasible alternatives to WHZ.

MUAC regression models indicated that higher MUAC, female sex, and stunting were associated with greater WHZ; older age and male sex were associated with significantly lower WHZ, suggesting that male children and children ages 24-59 months may be at greater risk for low weight-for-height. Age confounded the relationship between MUAC and WHZ. Model fit improved with the inclusion of adjustment covariates (Table S3A, Model 3, R^2^=0.4585), but less than half of the variability in WHZ was explained by its relationship with the other anthropometric indicators. MUACZ regression models showed a similar trend, as the age and sex adjustment inherent in MUACZ improved model fit (Table S3A, Model 4 R^2^=0.4171), but again failed to explain much of the variability in WHZ within these children.

While MUACZ and WHZ saw improved concordance (κ=0.385, fair concordance) compared to the MUAC and WHZ comparison, the sensitivity of 35.58% is still low. Eight children (0.6% of the sample) were identified as SAM by WHZ but normal by MUACZ, and 126 children (8.9% of the sample) were identified as MAM by WHZ but normal by MUACZ. This gap of 134 children, though smaller than the 195-child gap between MUAC and WHZ, still underestimates wasting prevalence. MUACZ thresholds may need further adjustment to ensure more wasted children receive treatment.

Further stratifying age into four categories (9-23 months, 24-35 months, 36-47 months, and 48-59 months) highlighted 2 years of age as a critical age point to recommend different MUAC cutoffs, in agreement with previous literature utilizing the same cutoff.[14,17,20,24] The 48–59 month age group also saw the highest concordance between MUACZ and WHZ, indicating potential improved agreement between MUACZ and WHZ around ages 4-5 years.

These results did not vary from midline to endline in the study, with only minor differences in concordance and adjusted MUAC thresholds. For children 9-23 months, the optimal MUAC threshold was 13.7cm at both midline and endline (Tables S14B and S6B, respectively). For children 24-59 months, the optimal MUAC threshold was 14.5cm at midline vs. 14.4cm at endline (Tables S14B and S6B, respectively). While recommended thresholds are slightly different, the underlying message is the same: raising thresholds for wasting may help identify and treat more truly wasted children in Somalia.

### 4.2 Programmatic Implications

Previous literature and this analysis raise several considerations for nutrition programs in humanitarian contexts. The 13 percentage point wasting prevalence difference using MUAC vs. WHZ in this analysis aligns with previous literature noting Somalia as an outlier due to larger differences in wasting prevalences using MUAC and WHZ compared to other countries.[15,21] MUAC as a sole diagnostic criterion failed to accurately identify 95% of WHZ-wasted children in this analysis and missed 25-80% of children in previous literature.[14,15,18–20] Program implementers and policy makers should be aware of the major limitations of using MUAC alone to diagnose wasting in CU5. If no action is taken, wasted children will remain untreated, keeping malnutrition high and vulnerable children at risk of mortality.

While rapidly incorporating WHZ into community screening may be an ideal way to diagnose wasting, the cost and complications of obtaining accurate weight and height measures in low-resource contexts may make this challenging. This study and previous evidence for updating wasting measurement guidelines prompt two alternative solutions for consideration: 1) Updating the MUAC threshold and stratifying by age at 24 months to align more closely with a WHZ of −2 SD or 2) Using the age-adjusted MUACZ as an alternative measurement.

Box 1 outlines the advantages, disadvantages, and implementation considerations of measures for diagnosing CU5 wasting during community screenings.

##### Comparison of Wasting Measurement Indicators

**Box 1. Comparison of Wasting Measurement Indicators**
| Measure | Advantages | Disadvantages | Implementation Needs/Considerations |
| --- | --- | --- | --- |
| *Current MUAC Threshold (MUAC <12.5cm)* | Currently in practice<br>No additional retraining/resources | Heavily underestimates true wasting burden<br>Misclassifies older children (24-59 months) at a higher rate | N/A |
| *Age-stratified MUAC threshold at 24 months* | Identifies more wasted children than current MUAC thresholds<br>Utilizes existing supply of MUAC tape | Minimal retraining for CHWs<br>Susceptible to inaccurate age data | Retraining CHWs<br>New policy requiring government and NGO support |
|  | Ease of measurement |  | Increased demand for wasting treatment infrastructure and supply |
| *MUACZ* | Identifies more wasted children than current MUAC thresholds<br>Ease of measurement | Lower sensitivity than other alternatives<br>Retraining for CHWs<br>Identifies additional children who may not be wasted using WHZ or MUAC<br>Susceptible to inaccurate age data | MUACZ tape supply<br>Retraining CHWs<br>New policy requiring government and NGO support<br>Increased demand for wasting treatment infrastructure and supply |
| *WHZ* | Identifies all wasted children<br>Most accurate wasting measurement<br>Aligns with current global recommendations | Costly supplies (compared to take)<br>Complicated measurements<br>Significant retraining for CHWs | Length boards and scale supply<br>Retraining CHWs<br>New policy requiring government and NGO support<br>Increased demand for wasting treatment infrastructure and supply |

Utilizing an updated and age-stratified MUAC threshold in this analysis correctly identified 75-80% of WHZ-wasted children, yielding a significant improvement over the current wasting threshold for MUAC. An age-stratified MUAC threshold accounts for clear differences in child growth before and after 24 months and ensures that children aged 24-59 months, who are especially vulnerable to this diagnostic misclassification, are more effectively screened for wasting. While the age-stratified MUAC threshold would ensure higher accuracy in wasting diagnosis, programmatic changes may complicate its implementation. Updating measurement guidelines would require retraining the health workforce and enrolling more children into wasting treatment, subsequently increasing the supplies, healthcare workers, and infrastructure needed for treatment. These requirements will impact the distribution and availability of supplies and increase funding needs for these programs. In settings without comprehensive vital registration, age-specific wasting guidelines may be also susceptible to inaccuracies in age data.

Alternatively, utilizing MUACZ to screen for wasting may offer similar benefits in diagnostic accuracy. In this analysis, MUACZ of −2 identified 36% of WHZ-wasted children and already performed far better than the current MUAC guidelines for wasting. This analysis suggested that even MUACZ thresholds should be adjusted to improve sensitivity, but updating wasting guidelines using MUACZ offers the same programmatic concerns for workforce, supply, and funding issues as the age-stratified MUAC thresholds.

The technical difficulties of anthropometry can reduce measurement accuracy and lead to diagnostic errors, especially in young children. If alternative measures for wasting diagnosis are to be considered, program implementers should take efforts to robustly train and retrain community health workers on best practices, while strengthening supervision and monitoring to maintain diagnostic accuracy.

It may be most feasible for Somalia to implement the solution of an increased MUAC threshold stratified by age at 24 months, rather than adopting new measurements. Yet, donors and nutrition stakeholders need to carefully consider how raising MUAC thresholds could impact health systems and health financing.

### 4.3 Study Limitations and Strengths

As with all evaluations, there are limitations to this work. Some known limitations include a smaller dataset, than many previous studies using global anthropometric data; however, our sample size was over n=1000, which is appropriately powered for these analyses. Stratified results must be interpreted with caution due to smaller sample sizes. The context changed from baseline to endline due to droughts, flooding, and displacement of study participants, which may have affected measurement rigor. Some participants were lost to follow up at endline, but the demographic distribution of the population remained consistent despite this loss. Vital registration data for child age in the study may be less accurate compared to other contexts. This study enrolled a healthy cohort at baseline (not wasted using MUAC) therefore wasting prevalence estimates may underestimate the true burden in these regions.

Our study does have considerable strengths. Our large sample size was well-powered for advanced statistical analyses and stratification. Our sample had a significantly higher wasting prevalence than the global average of 6.8%, and we produced timely wasting prevalences within 6 months of data collection. This study was conducted in a vulnerable context and directly identifies children who can benefit from more accurate screening measures for malnutrition. Our cluster randomized controlled trial may have more accurate and consistently measured anthropometric data than larger national survey datasets.

Future research into agreement between MUACZ and WHZ in global contexts should assess how various MUACZ thresholds improve sensitivity with WHZ. Future studies in Somalia should examine how well these recommended thresholds may perform across other regions and settings. Further studies should be conducted to assess the impact of adopting MUACZ and WHZ on workforce, supply, ease-of-use, and time requirements. Lastly, given the resource-intensive nature of implementing WHZ and MUACZ, additional costing analyses, such as Value for Money (VfM), should evaluate the financial implications and cost-effectiveness of scaling MUACZ or WHZ in low-resource settings.

## 5. Conclusion

Our results demonstrate poor concordance between MUAC and WHZ for diagnosing wasting among CU5 in two high wasting burden regions of Somalia. Although MUACZ had improved concordance, it still heavily underestimated WHZ wasting prevalence. WHZ is the gold standard for wasting diagnosis, but it remains an expensive and resource-intensive diagnostic technique. Age-stratified MUAC thresholds which identify a much larger proportion of WHZ-wasted children provide an alternative, more feasible diagnostic recommendation for community screening of malnutrition. For wasting screening in CU5 to be most effective, the global community must acknowledge the limitations of current anthropometric measures for wasting. Particularly international partners, donors, and government stakeholders in Somalia should consider this evidence and evaluate revision of wasting guidelines to treat and prevent future wasting in vulnerable children.

## Ethics Statement

Informed consent was obtained from all study participants. The study was conducted in accordance with the Declaration of Helsinki and approved by the Institutional Review Board of the Johns Hopkins Bloomberg School of Public Health (protocol code 24476, date of approval: May 17, 2023), the Ministry of Health and Human Services in Somalia (XAG/203/23, date of approval: April 26th, 2023), and the Save the Children International Ethics Review Committee.

## Disclosurement of Interest Statement

The authors declare no conflicts of interest.

## Data Availability Statement

This data was collected on a vulnerable population linked to a large scale cash assistance program in Somalia. The data may be made available upon request with permission from local study investigators.

## Authorship Contributions

Conceptualization and design of the work: SG, SW, KKA, SG, QK, SAM, AA, FMM, AMM, AM, MO, MBM, FL, MT, NA. Data analysis: SG, SW, KKA, SG, NA. Writing (original draft preparation) - SG, SW, KKA, SG, NA. Writing (review and editing) - SG, SW, KKA, SG, QK, SAM, AA, FMM, AMM, AM, MO, MBM, FL, MT, NA. Final approval of published submission - SG, SW, KKA, SG, QK, SAM, AA, FMM, AMM, AM, MO, MBM, FL, MT, NA. All authors have read and agree to the published version of this manuscript and agree to be accountable for all aspects of the work.

