## Supplemental Files for "Analyzing concordance between MUAC, MUACZ, and WHZ in diagnosing acute malnutrition among children under 5 in Somalia"

#### *Full Supplemental Files Table of Contents*

**Annex S1. Child Characteristics and Distributions - Endline**
Figure S1A. Distributions of Child Anthropometric Characteristics: Child Age, Height, MUAC, Weight
Figure S1B. Distributions of Child Anthropometric Characteristics: Z-Scores
Table S1C. Anthropometric Differences by Child Sex
Table S1D. Anthropometric Differences by Child Age (9-23 months vs. 24-59 months)
**Annex S2. Pearson's Correlation Figures - Endline**
Figure S2A. Pearson's Correlation between MUAC and WHZ
Figure S2B. Pearson's Correlation between MUAC and MUACZ
Figure S2C. Pearson's Correlation between MUACZ and WHZ
**Annex S3. Linear Regression Modelling Results - Endline**
Table S3A. Regression Modelling for Endline Sample
**Annex S4. Concordance Results - Endline**
Table S4A. Entire Sample (N=1408) Concordance Pairs: Wasted vs. Not Wasted
Table S4B. Entire Sample (N=1408) Concordance Pairs: Normal vs. MAM vs. SAM
Table S4C. Concordance Pairs Stratified by Region: Wasted vs. Not Wasted
Table S4D. Concordance Pairs Stratified by Age (9-23 months vs 24-59 months): Wasted vs. Not Wasted
Table S4E. Number of Children Diagnosed as Wasted Using MUAC and/or WHZ Criteria
Table S4F. Number of Children Diagnosed as Normal vs. MAM vs. SAM Using MUAC and/or WHZ Criteria
Table S4G. Number of Children Diagnosed as Wasted Using MUAC and/or MUACZ Criteria
Table S4H. Number of Children Diagnosed as Normal vs. MAM vs. SAM Using MUAC and/or MUACZ Criteria
Table S4I. Number of Children Diagnosed as Wasted Using MUACZ and/or WHZ Criteria
Table S4J. Number of Children Diagnosed as Normal vs. MAM vs. SAM Using MUACZ and/or WHZ Criteria
**Annex S5. Stratified Analysis: by Region - Endline**
Table S5A. Wasting Prevalence by Region
Table S5B. ROC Analysis by Region
**Annex S6. Stratified Analysis: by Child Age (9-23 mo. vs 24-59 mo.) - Endline**
Table S6A. Wasting Prevalence by Child Age
Table S6B. ROC Analysis by Child Age
**Table S7. Midline Analysis Demographics Table**
**Annex S8. Midline Analysis Child Characteristics and Distributions**
Figure S8A. Distributions of Child Anthropometric Characteristics: Child Age, Height, MUAC, Weight
Figure S8B. Distributions of Child Anthropometric Characteristics: Z-Scores
Table S8C. Anthropometric Differences by Child Sex
Table S8D. Anthropometric Differences by Child Age (9-23 months vs. 24-59 months)
**Annex S9. Midline Analysis Pearson's Correlation Figures**
Figure S9A. Pearson's Correlation between MUAC and WHZ
Figure S9B. Pearson's Correlation between MUAC and MUACZ
Figure S9C. Pearson's Correlation between MUACZ and WHZ
**Annex S10. Midline Analysis Linear Regression Modelling Results**
Table S10A. Regression Modelling for Endline Sample

**Annex S11. Midline Analysis Concordance Results**
Table S11A. Entire Sample Concordance Pairs: Wasted vs. Not Wasted
Table S11B. Entire Sample Concordance Pairs: Normal vs. MAM vs. SAM
Table S11C. Concordance Pairs Stratified by Region: Wasted vs. Not Wasted
Table S11D. Concordance Pairs Stratified by Age (9-23 months vs 24-59 months): Wasted vs. Not Wasted
Table S11E. Number of Children Diagnosed as Wasted Using MUAC and/or WHZ Criteria
Table S11F. Number of Children Diagnosed as Normal vs. MAM vs. SAM Using MUAC and/or WHZ Criteria
Table S11G. Number of Children Diagnosed as Wasted Using MUAC and/or MUACZ Criteria
Table S11H. Number of Children Diagnosed as Normal vs. MAM vs. SAM Using MUAC and/or MUACZ Criteria
Table S11I. Number of Children Diagnosed as Wasted Using MUACZ and/or WHZ Criteria
Table S11J. Number of Children Diagnosed as Normal vs. MAM vs. SAM Using MUACZ and/or WHZ Criteria
**Annex S12. Midline Full Sample ROC Analysis**
Table S12A. Current MUAC Thresholds, Sensitivity, Specificity, AUC – Midline
Table S12B. Ideal MUAC Thresholds, Sensitivity, Specificity, AUC
Table S12C. Ideal MUAC Thresholds, Sensitivity, Specificity, AUC
**Annex S13. Midline Stratified Analysis: by Region**
Table S13A. Wasting Prevalence by Region
Table S13B. ROC Analysis by Region
**Annex S14. Midline Stratified Analysis: by Child Age (9-23 mo. vs 24-59 mo.)**
Table S14A. Wasting Prevalence by Child Age
Table S14B. ROC Analysis by Child Age
**Annex S15. Description of Bay and Hiran Regions of Somalia**
**Annex S16. Detailed Description of CashPlus for Nutrition Study Protocol**
**Table S17. Study Outcome Definitions for Acute Malnutrition by Anthropometric Indicator**
**Annex S18. Additional Methods: Analysis Approach**

#### *Annex S1-S6 Table of Contents*

**Annex S1. Child Characteristics and Distributions - Endline**
Figure S1A. Distributions of Child Anthropometric Characteristics: Child Age, Height, MUAC, Weight
Figure S1B. Distributions of Child Anthropometric Characteristics: Z-Scores
Table S1C. Anthropometric Differences by Child Sex
Table S1D. Anthropometric Differences by Child Age (9-23 months vs. 24-59 months)
**Annex S2. Pearson's Correlation Figures - Endline**
Figure S2A. Pearson's Correlation between MUAC and WHZ
Figure S2B. Pearson's Correlation between MUAC and MUACZ
Figure S2C. Pearson's Correlation between MUACZ and WHZ
**Annex S3. Linear Regression Modelling Results - Endline**
Table S3A. Regression Modelling for Endline Sample
**Annex S4. Concordance Results - Endline**
Table S4A. Entire Sample (N=1408) Concordance Pairs: Wasted vs. Not Wasted
Table S4B. Entire Sample (N=1408) Concordance Pairs: Normal vs. MAM vs. SAM
Table S4C. Concordance Pairs Stratified by Region: Wasted vs. Not Wasted
Table S4D. Concordance Pairs Stratified by Age (9-23 months vs 24-59 months): Wasted vs. Not Wasted
Table S4E. Number of Children Diagnosed as Wasted Using MUAC and/or WHZ Criteria
Table S4F. Number of Children Diagnosed as Normal vs. MAM vs. SAM Using MUAC and/or WHZ Criteria
Table S4G. Number of Children Diagnosed as Wasted Using MUAC and/or MUACZ Criteria
Table S4H. Number of Children Diagnosed as Normal vs. MAM vs. SAM Using MUAC and/or MUACZ Criteria
Table S4I. Number of Children Diagnosed as Wasted Using MUACZ and/or WHZ Criteria
Table S4J. Number of Children Diagnosed as Normal vs. MAM vs. SAM Using MUACZ and/or WHZ Criteria
**Annex S5. Stratified Analysis: by Region - Endline**
Table S5A. Wasting Prevalence by Region
Table S5B. ROC Analysis by Region
**Annex S6. Stratified Analysis: by Child Age (9-23 mo. vs 24-59 mo.) - Endline**
Table S6A. Wasting Prevalence by Child Age
Table S6B. ROC Analysis by Child Age

**Annex S1. Child Characteristics and Distributions - Endline**

**Figure S1A. Distributions of Child Anthropometric Characteristics: Child Age, Height, MUAC, Weight**

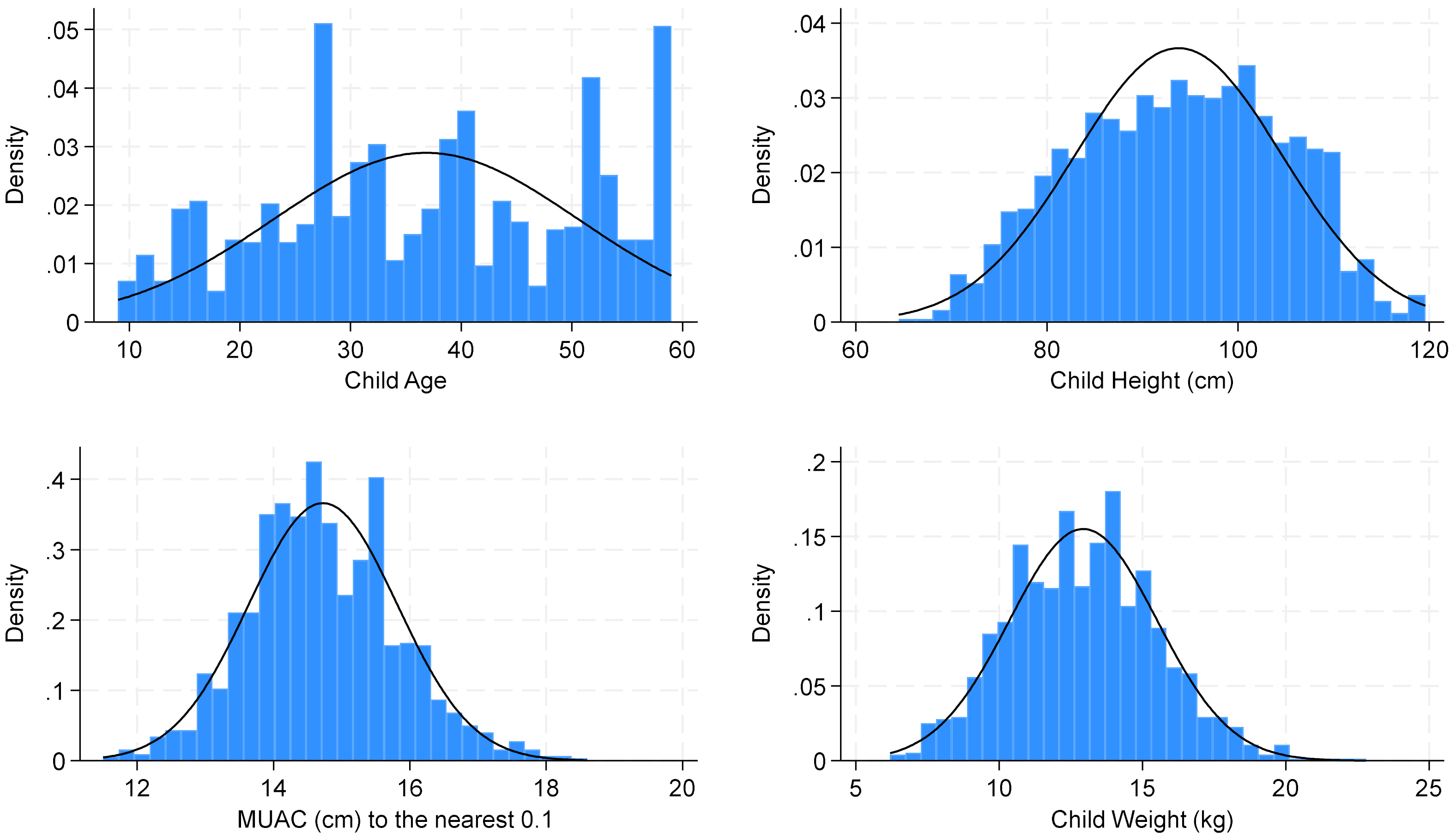

**Figure S1B. Distributions of Child Anthropometric Characteristics: Z-Scores**

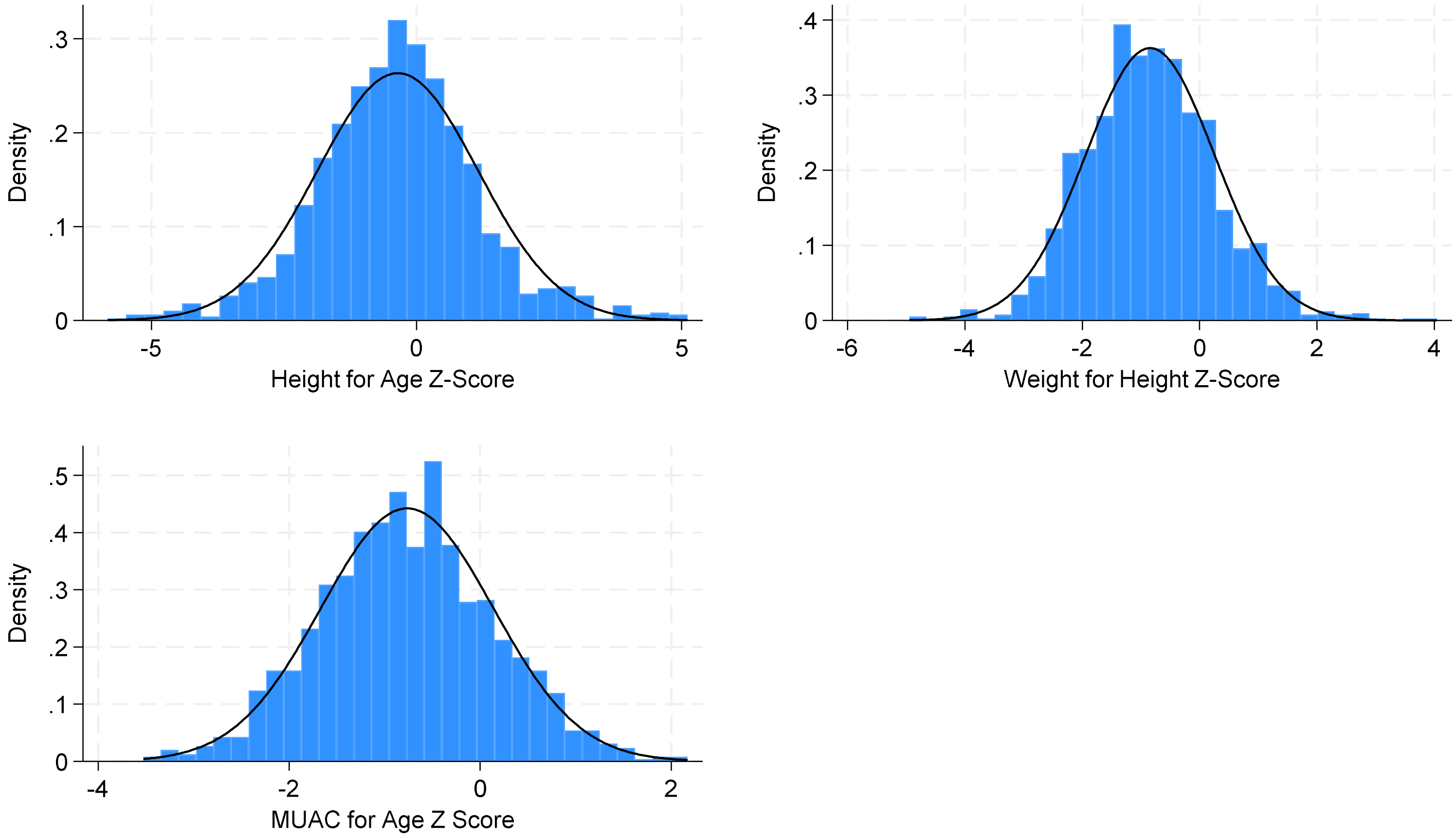

**Table S1C. Anthropometric Differences by Child Sex**

| Anthropometric Measurement | Male (n=687) | Female (n=700) | p-value |
|---|---|---|---|
| Wasting Prevalence by MUAC | 1.72% (n=12) | 1.27% (n=9) | 0.489 |
| Mean MUAC (cm) | 14.72cm | 14.74cm | 0.748 |
| Wasting Prevalence by WHZ | 16.88% (n=118) | 12.69% (n=90) | 0.027* |
| Mean WHZ | -0.93 SD | -0.76 SD | 0.003* |

*p<0.05.

**Table S1D. Anthropometric Differences by Child Age (9-23 months vs. 24-59 months)**

| Anthropometric Measurement | 9-23 months (n=270) | 24-59 months (n=1,138) | p-value |
|---|---|---|---|
| Wasting Prevalence by MUAC | 4.07% (n=11) | 0.88% (n=10) | <0.001* |
| Mean MUAC (cm) | 14.18cm | 14.86cm | <0.001* |
| Wasting Prevalence by WHZ | 12.22% (n=33) | 15.38% (n=175) | 0.189 |
| Mean WHZ | -0.65 SD | -0.89 SD | 0.002* |

*p<0.05.

#### Annex S2. Pearson's Correlation Figures - Endline

##### Figure S2A. Pearson's Correlation between MUAC and WHZ: ρ=0.5501

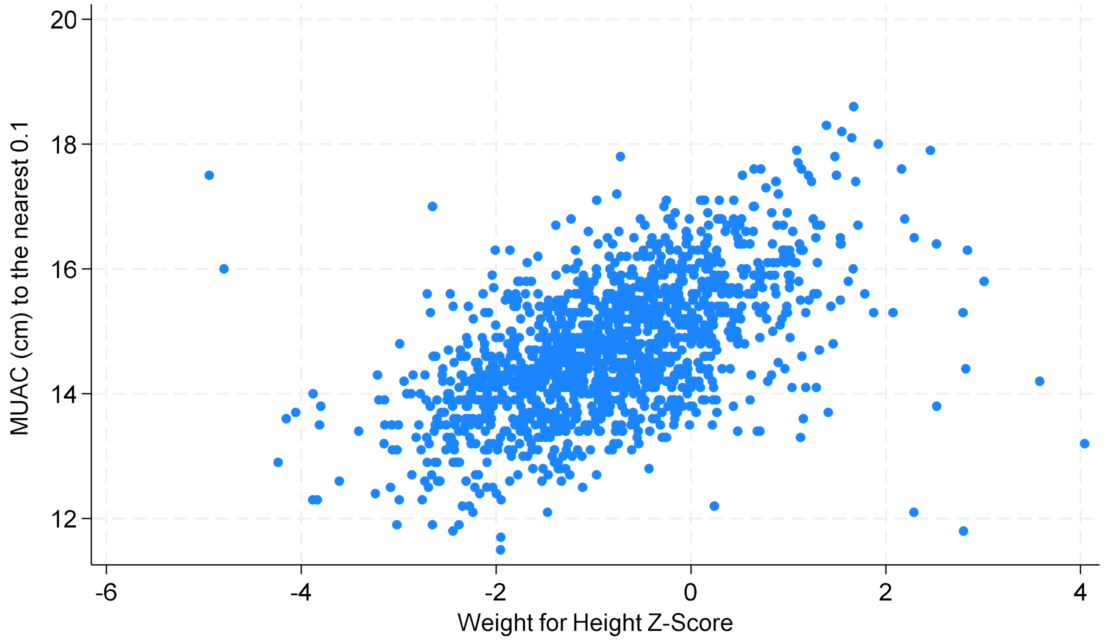

**Figure S2B. Pearson's Correlation between MUAC and MUACZ: ρ=0.8455**

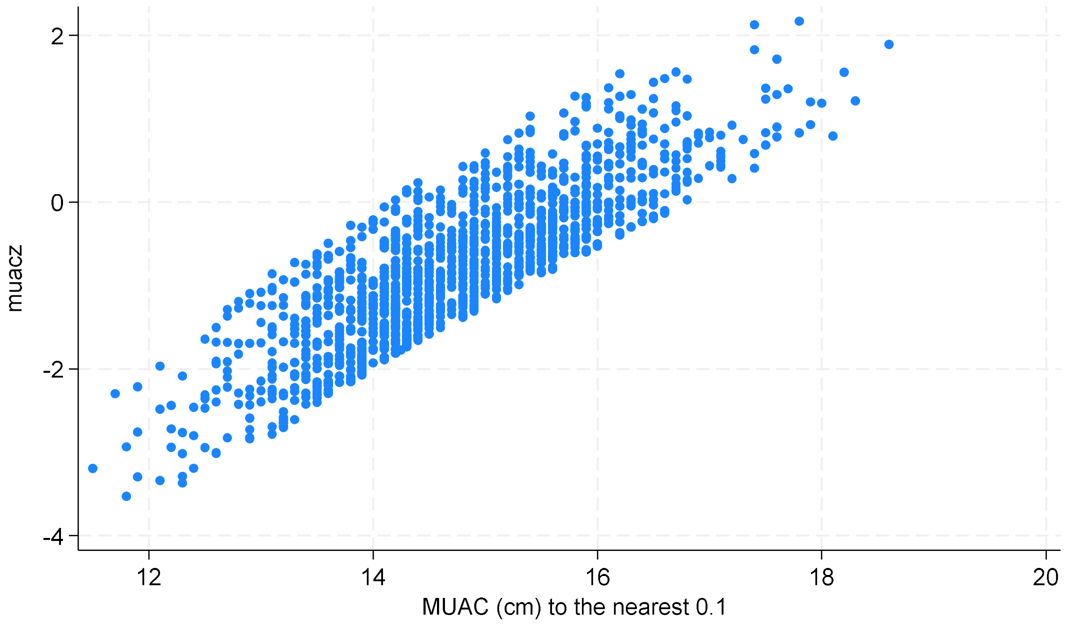

**Figure S2C. Pearson's Correlation between MUACZ and WHZ: ρ=0.6458**

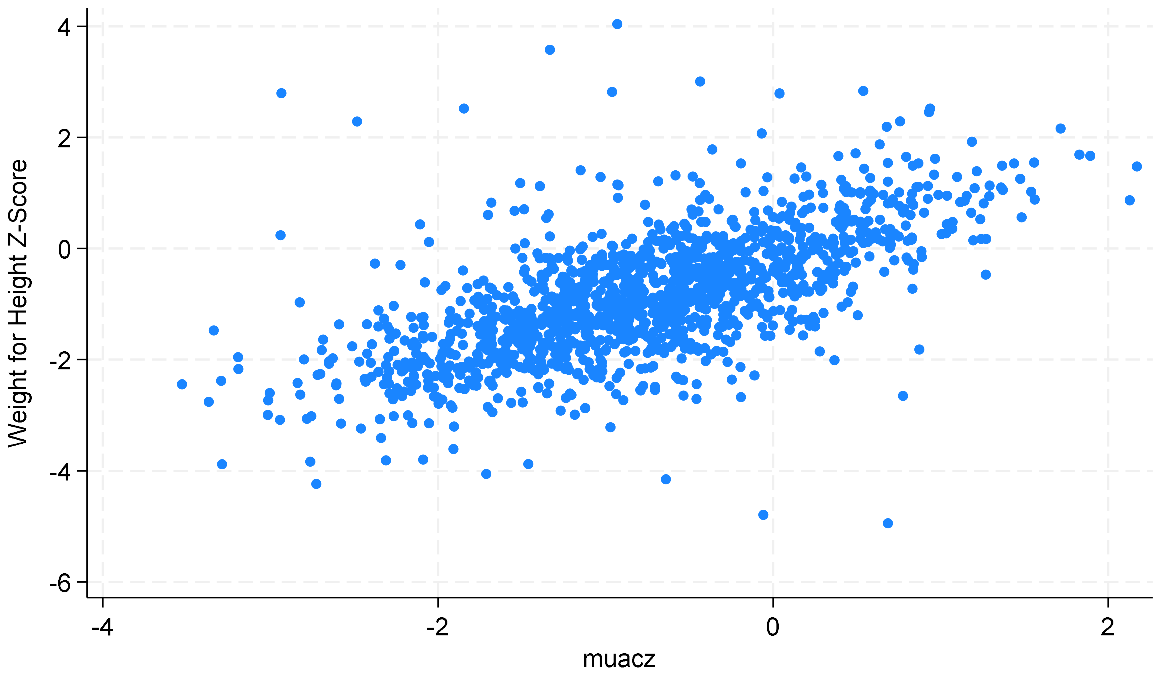

#### Annex S3. Regression Modelling – Endline

##### Table S3A. Regression Modelling for Endline Sample

| | **Model 1** β (95% CI) | **Model 2** β (95% CI) | **Model 3** β (95% CI) | **Model 4** β (95% CI) | **Model 5** β (95% CI) | **Model 6** β (95% CI) | **Model 7** β (95% CI) | **Model 8** β (95% CI) |
|---|---|---|---|---|---|---|---|---|
| MUAC | 0.56*** (0.51, 0.60) | 0.65*** (0.61, 0.69) | 0.67*** (0.63, 0.72) | 0.68*** (0.63, 0.72) | 0.66*** (0.62, 0.77) | - | - | 0.67*** (0.63,0.71) |
| Age (continuous) | - | -0.03*** (-0.03, -0.03) | -0.03*** (-0.03, -0.03) | -0.03*** (-0.03, -0.02) | -0.03*** (-0.03, -0.02) | - | 0.0002 (-0.003, 0.003) | - |
| Sex (Female) | - | - | 0.16*** (0.07, 0.24) | 0.21* (0.05, 0.37) | 0.15*** (0.07, 0.24) | - | 0.07 (-0.02, 0.15) | 0.16*** (0.07, 0.24) |
| Stunting (Stunted vs. not stunted) | - | - | 0.65*** (0.51, 0.78) | 0.65*** (0.52, 0.78) | 0.60** (0.47, 0.74) | - | 0.66*** (0.53, 0.80) | 0.65*** (0.51, 0.78) |
| Age * Sex | - | - | - | -0.06 (-0.24, 0.11) | - | - | - | - |
| Region (Bay vs Hiran) | - | - | - | - | 0.14** (0.04, 0.23) | - | - | - |
| MUACZ | - | - | - | - | - | 0.79*** (0.74, 0.84) | 0.81*** (0.76, 0.86) | - |
| Age (reference category 9-23 months) | - | - | - | - | - | - | - | - |
| 24-35 months | - | - | - | - | - | - | - | -0.38*** (-0.51, -0.26) |
| 36-47 months | - | - | - | - | - | - | - | -0.69*** (-0.82, -0.55) |
| 48-59 months | - | - | - | - | - | - | - | -1.04*** (-1.17, -0.91) |
| **Model $R^2$** | **0.3026** | **0.4176** | **0.4585** | **0.4587** | **0.4618** | **0.4171** | **0.4554** | **0.4489** |

Note: MUAC and MUACZ were modelled as continuous variables. Sex, stunting, and region were modelled as dichotomous variables. Age was modelled as both a continuous variable and a categorical variable, depending on the model. $^{*}p<0.05$. $^{**}p<0.01$. $^{***}p<0.001$.

Note on Model 8: Linear combinations for this regression model indicated that all categories were statistically significant from each other and all p<0.001.

### Annex S4. Tables with Concordance Pairs - Endline

**Table S4A. Entire Sample (N=1408) Concordance Pairs: Wasted vs. Not Wasted**

| Pair of Wasting Indicators | Kappa | Strength of Concordance |
|---|---|---|
| MUAC & WHZ | 0.0889 | None-slight |
| MUAC & MUACZ | 0.2650 | Fair |
| MUACZ & WHZ | 0.3847 | Fair (almost moderate) |

**Table S4B. Entire Sample (N=1408) Concordance Pairs: Normal vs. MAM vs. SAM**

| Pair of Wasting Indicators | Kappa | Strength of Concordance |
|---|---|---|
| MUAC & WHZ | 0.0724 | None-slight |
| MUAC & MUACZ | 0.2077 | Fair |
| MUACZ & WHZ | 0.3169 | Fair |

**Table S4C. Concordance Pairs Stratified by Region: Wasted vs. Not Wasted**

| Pair of Wasting Indicators | Bay Kappa | Hiran Kappa | Total |
|---|---|---|---|
| MUAC & WHZ | 0.0931 | 0.0846 | 0.0889 |
| MUAC & MUACZ | 0.3209 | 0.2435 | 0.2650 |
| MUACZ & WHZ | 0.2928 | 0.1761 | 0.3847 |

**Table S4D. Concordance Pairs Stratified by Age (9-23 months vs 24-59 months): Wasted vs. Not Wasted**

| Pair of Wasting Indicators | 9-23 months Kappa | 24-59 months Kappa | Total |
|---|---|---|---|
| MUAC & WHZ | 0.2254 | 0.06 | 0.0889 |
| MUAC & MUACZ | 0.7578 | 0.1604 | 0.2650 |
| MUACZ & WHZ | 0.2782 | 0.4025 | 0.3847 |

**Table S4E. Number of Children Diagnosed as Wasted Using MUAC and/or WHZ Criteria**

| Child Wasting by MUAC | | Child Wasting by WHZ: Not wasted | Child Wasting by WHZ: Wasted | **Total** |
|---|---|---|---|---|
| | Not wasted | 1192 | 195 | 1387 |
| | Wasted | 8 | 13 | 21 |
| | **Total** | 1200 | 208 | 1408 |

**Table S4F. Number of Children Diagnosed as Normal vs. MAM vs. SAM Using MUAC and/or WHZ Criteria**

| Child Wasting by MUAC | | Child Wasting by WHZ: SAM | Child Wasting by WHZ: MAM | Child Wasting by WHZ: Normal | **Total** |
|---|---|---|---|---|---|
| | SAM | 0 | 0 | 0 | 0 |
| | MAM | 4 | 9 | 8 | 21 |
| | Normal | 20 | 185 | 1192 | 1387 |
| | **Total** | 24 | 184 | 1200 | 1408 |

*Note: No children were diagnosed as SAM by MUAC (<11.5cm)

**Table S4G. Number of Children Diagnosed as Wasted Using MUAC and/or MUACZ Criteria**

| | | Child Wasting by MUACZ | | |
|---|---|---|---|---|
| | | Not wasted | Wasted | **Total** |
| **Child Wasting by MUAC** | Not wasted | 1287 | 100 | 1387 |
| | Wasted | 1 | 20 | 21 |
| | **Total** | 1288 | 120 | 1408 |

**Table S4H. Number of Children Diagnosed as Normal vs. MAM vs. SAM Using MUAC and/or MUACZ Criteria**

| | | Child Wasting by MUACZ | | | |
|---|---|---|---|---|---|
| | | SAM | MAM | Normal | **Total** |
| **Child Wasting by MUAC** | SAM | 0 | 0 | 0 | 0 |
| | MAM | 8 | 12 | 1 | 21 |
| | Normal | 2 | 98 | 1287 | 1387 |
| | **Total** | 10 | 110 | 1288 | 1408 |

**Table S4I. Number of Children Diagnosed as Wasted Using MUACZ and/or WHZ Criteria**

| | | Child Wasting by WHZ | | |
|---|---|---|---|---|
| | | Not wasted | Wasted | **Total** |
| **Child Wasting by MUACZ** | Not wasted | 1154 | 134 | 1288 |
| | Wasted | 46 | 74 | 120 |
| | **Total** | 1200 | 208 | 1408 |

**Table S4J. Number of Children Diagnosed as Normal vs. MAM vs. SAM Using MUACZ and/or WHZ Criteria**

| | | Child Wasting by WHZ | | | |
|---|---|---|---|---|---|
| | | SAM | MAM | Normal | **Total** |
| **Child Wasting by MUACZ** | SAM | 1 | 7 | 2 | 10 |
| | MAM | 15 | 51 | 44 | 110 |
| | Normal | 8 | 126 | 1154 | 1288 |
| | **Total** | 24 | 184 | 1200 | 1408 |

#### Annex S5. Stratified Analysis by Region (Bay vs. Hiran) - Endline

##### Table S5A. Wasting Prevalence by Region (Bay vs. Hiran)

| **Indicator** | **Bay** (n=614) | **Hiran** (n=794) | **p-values** |
|---|---|---|---|
| MUAC | **1.14%** (n=7)<br>(95% CI: 0.54%, 2.38%) | **1.76%** (n=14)<br>(95% CI: 1.05%, 2.96%) | 0.339 |
| WHZ | **7.65%** (n=47)<br>(95% CI: 5.80%, 10.05%) | **20.28%** (n=161)<br>(95% CI: 17.62%, 23.22%) | <0.001* |
| Oedema | **0.33%** (n=2)<br>(95% CI: 0.08%, 1.30%) | **0.25%** (n=2)<br>(95% CI: 0.06%, 1.00%) | 0.796 |
| WHO Definition Wasted:<br>by MUAC or WHZ or Oedema | **8.47%** (n=52)<br>(95% CI: 6.51%, 10.95%) | **20.91%** (n=166)<br>(95% CI: 18.21%, 23.88%) | <0.001* |
| MUACZ | **4.72%** (n=29)<br>(95% CI: 3.30%, 6.72%) | **11.46%** (n=91)<br>(95% CI: 9.42%, 13.87%) | <0.001* |
| Wasting by all 4 Measures:<br>by MUAC, WHZ, Oedema, or MUACZ | **10.42%** (n=64)<br>(95% CI: 8.24%, 13.11%) | **24.18%** (n=192)<br>(95% CI: 21.35%, 27.29%) | <0.001* |

*p<0.05

##### Table S5B. ROC Analysis by Region (Bay vs. Hiran)

| **ROC Analysis Results** | **Bay** | **Hiran** |
|---|---|---|
| Ideal MUAC Threshold | 14.4cm | 14.3cm |
| MUAC AUC | 0.8012 | 0.7123 |
| MUAC Sensitivity | 87.23% | 74.53% |
| MUAC Specificity | 73.02% | 67.93% |
| Ideal MUACZ Threshold | -0.9 SD | -1.1 SD |
| MUACZ AUC | 0.7800 | 0.7308 |
| MUACZ Sensitivity | 85.11% | 79.50% |
| MUACZ Specificity | 70.90% | 66.67% |

Note: There was minimal difference in the ideal MUAC by region, but the sensitivities, specificities, and AUCs are very different, but this is likely due to differences in age or stunting prevalence by region. Similarly to MUAC, there was minimal difference in the ideal MUACZ by region, but the sensitivities, specificities, and AUCs are different.

#### Annex S6. Stratified Analysis by Child Age (9-23 months vs 24-59 months) - Endline

##### Table S6A. Wasting Prevalences by Child Age (9-23 months vs 24-59 months)

| **Indicator** | **9-23 months** (n=273) | **24-59 months** (n=1,138) | **p-values** |
|---|---|---|---|
| MUAC | **4.07%** (n=11)<br>(95% CI: 2.26%, 7.22%) | **0.88%** (n=10)<br>(95% CI: 0.47%, 1.63%) | <0.001* |
| WHZ | **12.22%** (n=33)<br>(95% CI: 8.81%, 16.72%) | **15.38%** (n=175)<br>(95% CI: 13.39%, 17.59%) | 0.189 |
| Oedema | **0.37%** (n=1)<br>(95% CI: 0.05%, 2.6%%) | **0.26%** (n=3)<br>(95% CI: 0.08%, 0.82%) | 0.767 |
| WHO Definition Wasted: by MUAC **or** WHZ **or** Oedema | **14.07%** (n=38)<br>(95% CI: 10.40%, 18.78%) | **15.82%** (n=180)<br>(95% CI: 13.81%, 18.06%) | 0.477 |
| MUACZ | **5.56%** (n=15)<br>(95% CI: 3.37%, 9.03%) | **9.23%** (n=105)<br>(95% CI: 7.68%, 11.05%) | 0.052 |
| Wasting by all 4 Measures: (MUAC, WHZ, Oedema, MUACZ) | **14.81%** (n=40)<br>(95% CI: 11.04%, 19.59%) | **18.98%** (n=216)<br>(95% CI: 16.80%, 21.37%) | 0.111 |

*p<0.05 **Note:** There was a borderline significant difference in wasting prevalence by MUACZ between the age groups. These differences should be interpreted with caution due to the difference in sample sizes between the two age groups and small size of the 9-23 months old group.

##### Table S6B. ROC Analysis by Child Age (9-23 months vs 24-59 months)

| **ROC Analysis Results** | **9-23 months** | **24.59 months** |
|---|---|---|
| Ideal MUAC Threshold | 13.7cm | 14.4cm |
| MUAC AUC | 0.7762 | 0.7492 |
| MUAC Sensitivity | 81.82% | 76.00% |
| MUAC Specificity | 73.42% | 73.83% |
| Ideal MUACZ Threshold | -0.9 SD | -1 SD |
| MUACZ AUC | 0.8019 | 0.7523 |
| MUACZ Sensitivity | 84.85% | 85.14% |
| MUACZ Specificity | 75.53% | 65.32% |

#### *Table of Contents*

**Table S7. Midline Analysis Demographics Table**
**Annex S8. Midline Analysis Child Characteristics and Distributions**
Figure S8A. Distributions of Child Anthropometric Characteristics: Child Age, Height, MUAC, Weight
Figure S8B. Distributions of Child Anthropometric Characteristics: Z-Scores
Table S8C. Anthropometric Differences by Child Sex
Table S8D. Anthropometric Differences by Child Age (9-23 months vs. 24-59 months)
**Annex S9. Midline Analysis Pearson's Correlation Figures**
Figure S9A. Pearson's Correlation between MUAC and WHZ
Figure S9B. Pearson's Correlation between MUAC and MUACZ
Figure S9C. Pearson's Correlation between MUACZ and WHZ
**Annex S10. Midline Analysis Linear Regression Modelling Results**
Table S10A. Regression Modelling for Endline Sample
**Annex S11. Midline Analysis Concordance Results**
Table S11A. Entire Sample Concordance Pairs: Wasted vs. Not Wasted
Table S11B. Entire Sample Concordance Pairs: Normal vs. MAM vs. SAM
Table S11C. Concordance Pairs Stratified by Region: Wasted vs. Not Wasted
Table S11D. Concordance Pairs Stratified by Age (9-23 months vs 24-59 months): Wasted vs. Not Wasted
Table S11E. Number of Children Diagnosed as Wasted Using MUAC and/or WHZ Criteria
Table S11F. Number of Children Diagnosed as Normal vs. MAM vs. SAM Using MUAC and/or WHZ Criteria
Table S11G. Number of Children Diagnosed as Wasted Using MUAC and/or MUACZ Criteria
Table S11H. Number of Children Diagnosed as Normal vs. MAM vs. SAM Using MUAC and/or MUACZ Criteria
Table S11I. Number of Children Diagnosed as Wasted Using MUACZ and/or WHZ Criteria
Table S11J. Number of Children Diagnosed as Normal vs. MAM vs. SAM Using MUACZ and/or WHZ Criteria
**Annex S12. Midline Full Sample ROC Analysis**
Table S12A. Current MUAC Thresholds, Sensitivity, Specificity, AUC – Midline
Table S12B. Ideal MUAC Thresholds, Sensitivity, Specificity, AUC
Table S12C. Ideal MUAC Thresholds, Sensitivity, Specificity, AUC
**Annex S13. Midline Stratified Analysis: by Region**
Table S13A. Wasting Prevalence by Region
Table S13B. ROC Analysis by Region
**Annex S14. Midline Stratified Analysis: by Child Age (9-23 mo. vs 24-59 mo.)**
Table S14A. Wasting Prevalence by Child Age
Table S14B. ROC Analysis by Child Age

#### Table S7. Midline Analysis Demographics Table

| Sample Characteristics (N=1,482) | | | | |
|---|---|---|---|---|
| **Demographic Characteristics** | **Total Sample** | Hiran (n=869) | Bay (n=613) | p values |
| Child Sex, n (%) | | | | |
| Male | 730 (49.26%) | 448 (51.55%) | 282 (46.00%) | 0.035 |
| Female | 752 (50.74%) | 421 (48.45%) | 331 (54.00%) | |
| Child Age, mean (SD) | 35.78 (13.37) | 37.49 (13.59) | 33.35 (12.66) | <0.001 |
| 9-23 months (n, %) | 303 (20.45%) | 162 (18.64%) | 141 (23.00%) | 0.040 |
| 23-59 months (n, %) | 1,179 (79.55%) | 707 (81.36%) | 472 (77.00%) | |
| Child Age - 4 categories* | | | | |
| 9-23 months (n, %) | 303 (20.45%) | 162 (18.64%) | 141 (23.00%) | <0.001 |
| 24-35 months (n, %) | 431 (29.08%) | 215 (24.74%) | 216 (35.24%) | |
| 36-47 months (n, %) | 374 (25.24%) | 230 (26.47%) | 144 (23.49%) | |
| 48-59 months (n, %) | 374 (25.24%) | 262 (30.15%) | 112 (18.27%) | |
| **Anthropometric Data,** mean (SD) | | | | |
| Child MUAC (cm) | 14.62 (1.06) | 14.48 (0.99) | 14.85 (1.12) | <0.001 |
| Weight (kg) | 12.44 (2.44) | 12.77 (2.47) | 11.96 (2.32) | <0.001 |
| Height (cm) | 91.30 (10.90) | 93.86 (10.75) | 87.66 (10.03) | <0.001 |
| **Z Scores**, mean (SD) | | | | |
| WHZ | -0.74 (1.11) | -1.00 (1.09) | -0.36 (1.04) | <0.001 |
| HAZ | -0.89 (1.43) | -0.49 (1.29) | -1.44 (1.44) | <0.001 |
| MUACZ | -0.81 (0.88) | -0.99 (0.83) | -0.55 (0.88) | <0.001 |
| **Stunting Prevalence,** n (%) | | | | |
| Not Stunted | 1,185 (79.96%) | 773 (88.95%) | 412 (67.21%) | <0.001 |
| Stunted | 297 (20.04%) | 96 (11.05%) | 201 (32.79%) | |
| **Wasting Prevalences by Indicator,** n (%) [95% CI] | | | | |
| MUAC | 18 (1.21%) [7.66-19.20%] | 6 (0.98%) [0.20%, 1.76%] | 12 (1.38%) [0.61%, 2.16%] | 0.486 |
| WHZ | 189 (12.75%) [11.11-14.55%] | 32 (5.22%) [3.46- 6.98%] | 157 (18.01%) [15.51%, 20.62%] | <0.001* |
| Edema | 14 (0.94%) [0.56-1.59%] | 12 (1.38%) [0.79-2.42%] | 2 (0.33%) [0.08-1.30%] | 0.039 |
| 2013 WHO Wasting Guidelines: by MUAC or WHZ or Edema | 209 (14.10%) [12.42-15.97%] | 36 (5.87%) [4.01-7.73%] | 173 (19.91%) [17.25-22.56%] | <0.001* |
| MUACZ | 123 (8.30%) | 28 (4.57%) [ 2.91-6.22%] | 95 (10.93%) [8.86-13.01%] | <0.001* |
| Wasting by all 4 Measures: (MUAC, WHZ, Oedema, or MUACZ) | [7.00-9.82%] | 51 (8.32%) [6.13-10.51%] | 203 (23.36%) [20.55-26.17%] | <0.001* |

#### Annex S8. Midline Analysis Child Characteristics and Distributions

**Figure S8A. Distributions of Child Anthropometric Characteristics: Child Age, Height, MUAC, Weight**

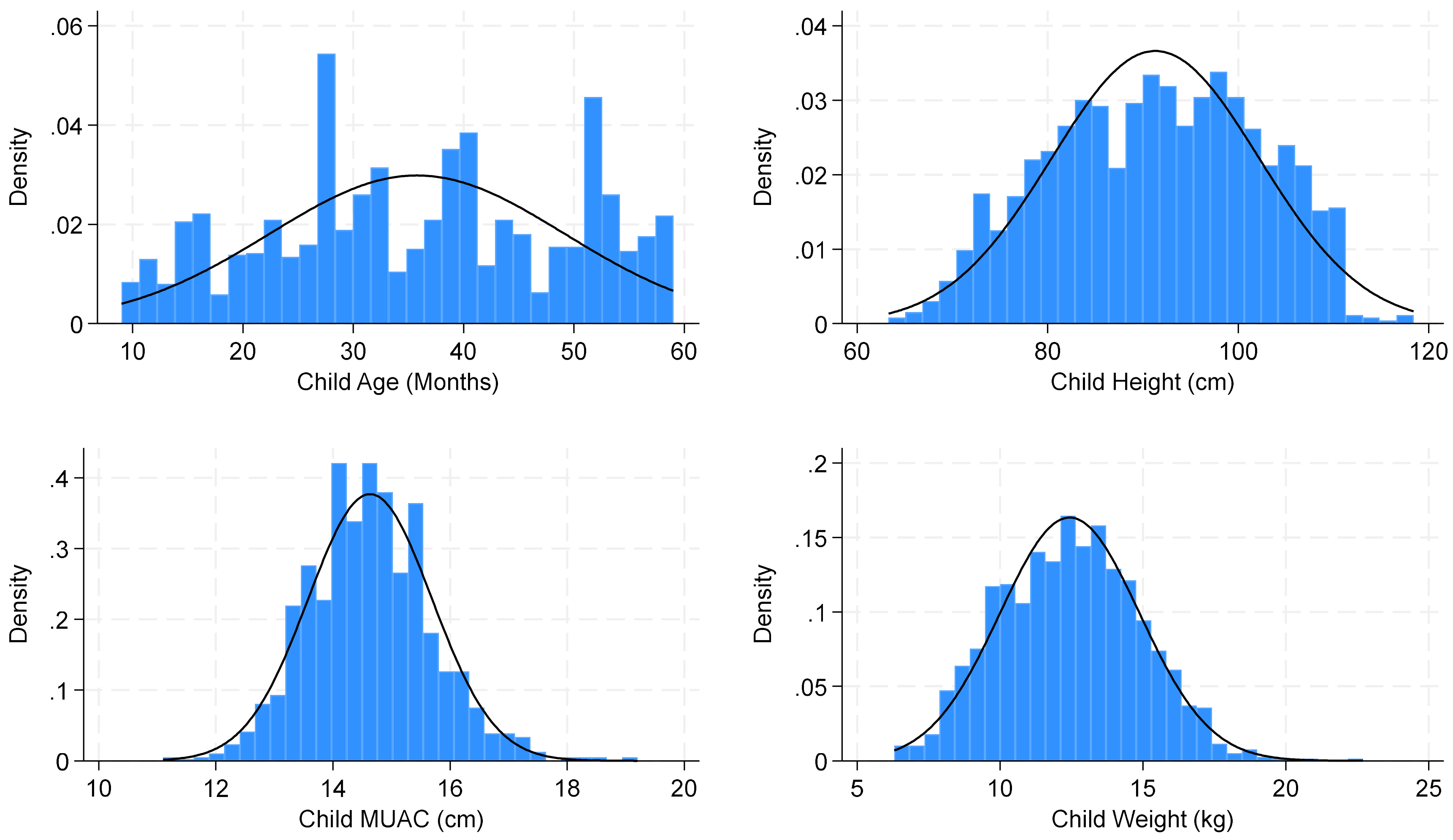

**Figure S8B. Distributions of Child Anthropometric Characteristics: Z-Scores**

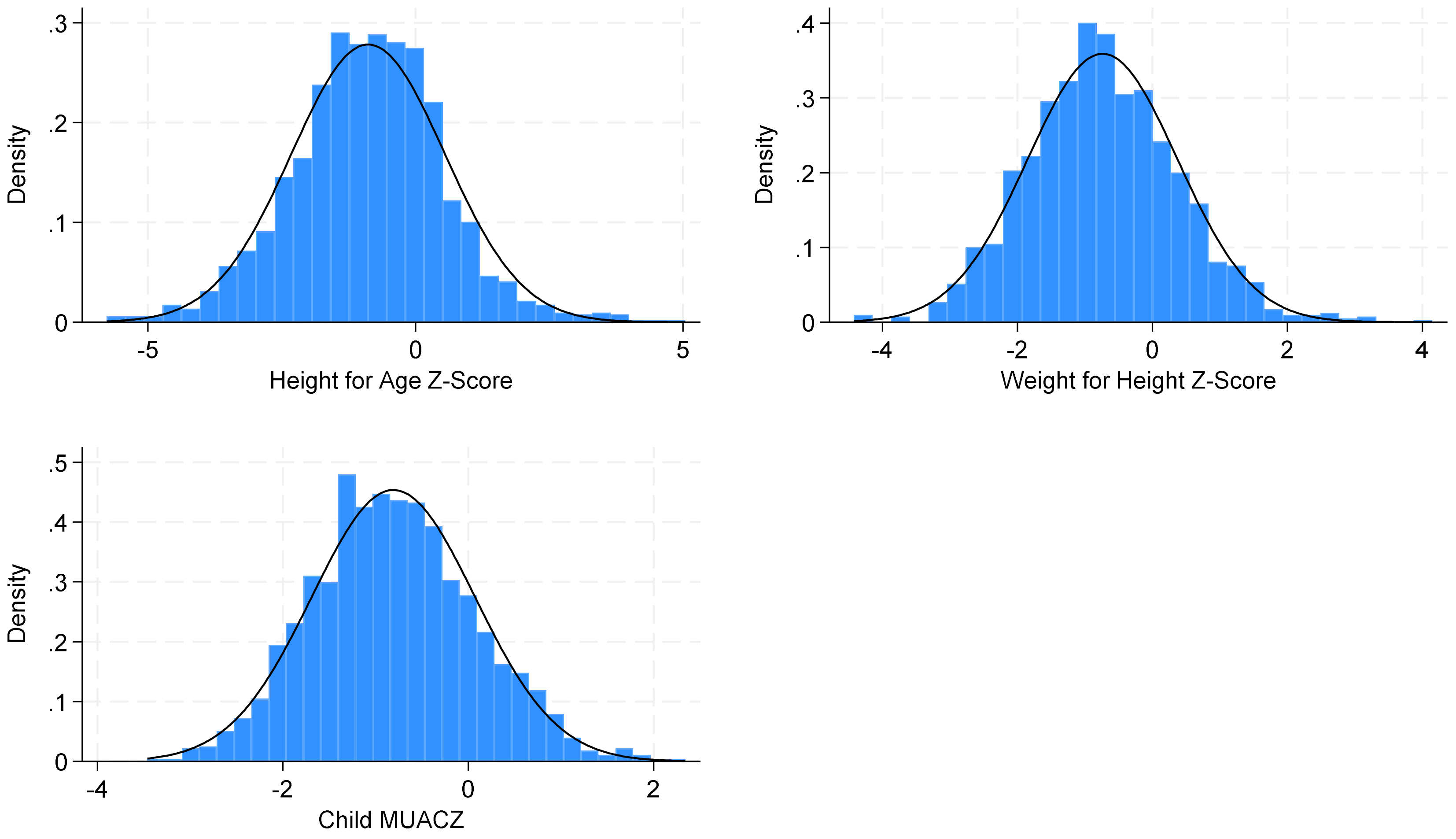

**Table S8C. Anthropometric Differences by Child Sex**

| Anthropometric Measurement | Male (n=730) | Female (n=752) | p-value |
|---|---|---|---|
| Wasting Prevalence by MUAC | 0.55% (n=4) | 1.86% (n=14) | 0.021* |
| Mean MUAC (cm) | 14.61cm | 14.66cm | 0.361 |
| Wasting Prevalence by WHZ | 14.93% (n=109) | 10.64% (n=80) | 0.013* |
| Mean WHZ | -0.83 SD | -0.65 SD | 0.001* |

*p<0.05.

**Table S8D. Anthropometric Differences by Child Age (9-23 months vs. 24-59 months)**

| Anthropometric Measurement | 9-23 months (n=303) | 24-59 months (n=1,179) | p-value |
|---|---|---|---|
| Wasting Prevalence by MUAC | 4.29% (n=13) | 0.42% (n=5) | <0.001* |
| Mean MUAC (cm) | 14.07cm | 14.78cm | <0.001* |
| Wasting Prevalence by WHZ | 10.56% (n=32) | 13.32% (n=157) | 0.200 |
| Mean WHZ | -0.49 SD | -0.80 SD | <0.001* |

*p<0.05.

#### Annex S9. Midline Analysis Pearson's Correlation Figures

**Figure S9A. Pearson's Correlation between MUAC and WHZ: ρ=0.5617**

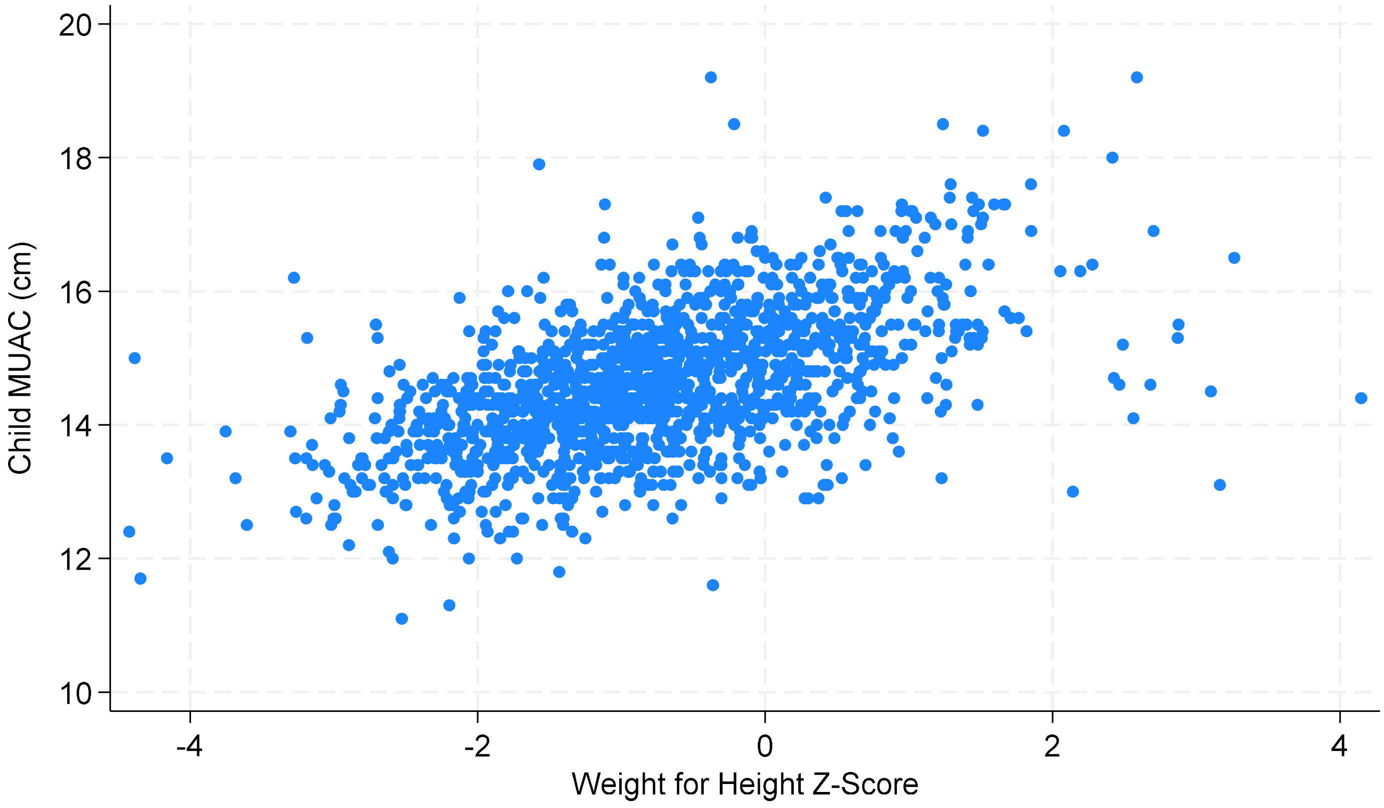

**Figure S9B. Pearson's Correlation between MUAC and MUACZ: ρ=0.8458**

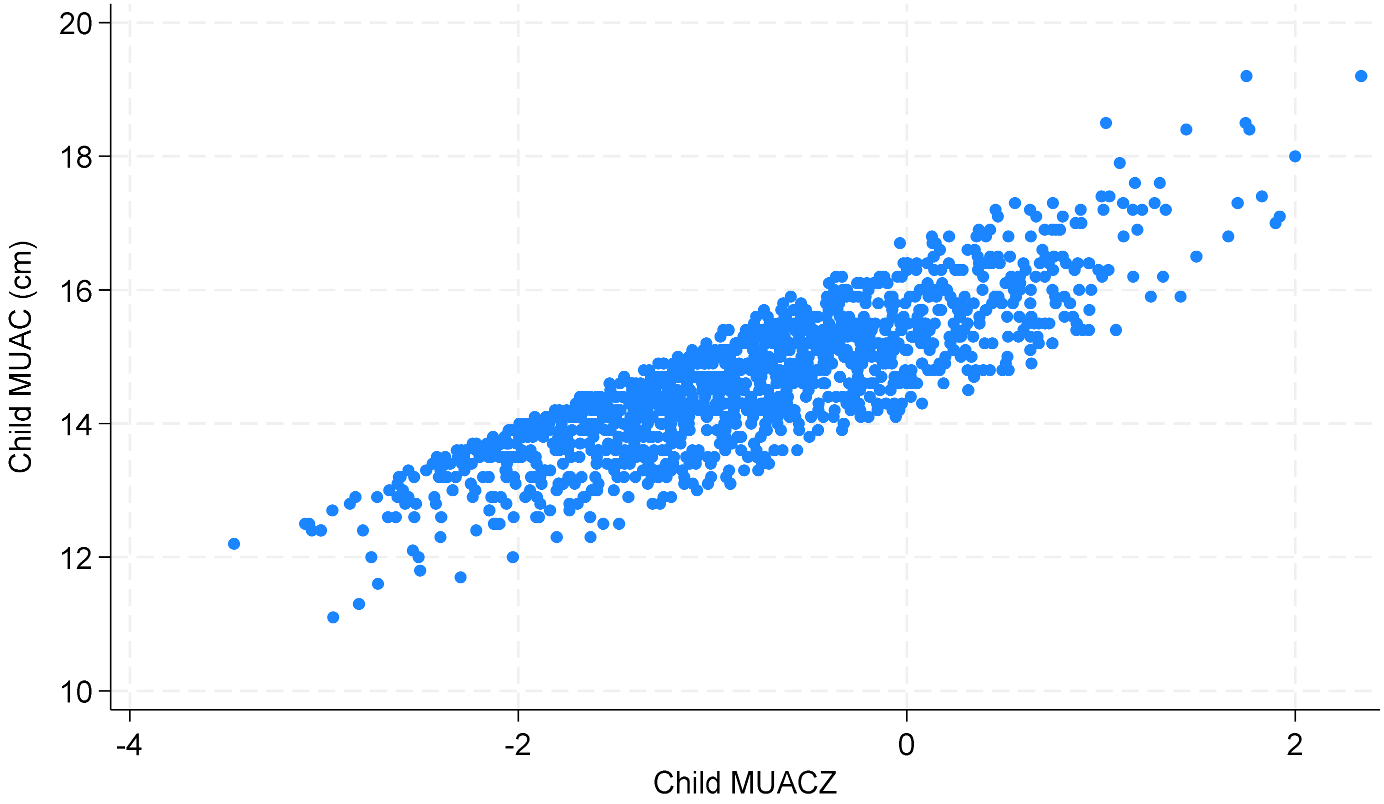

**Figure S9C. Pearson's Correlation between MUACZ and WHZ: ρ=0.6898**

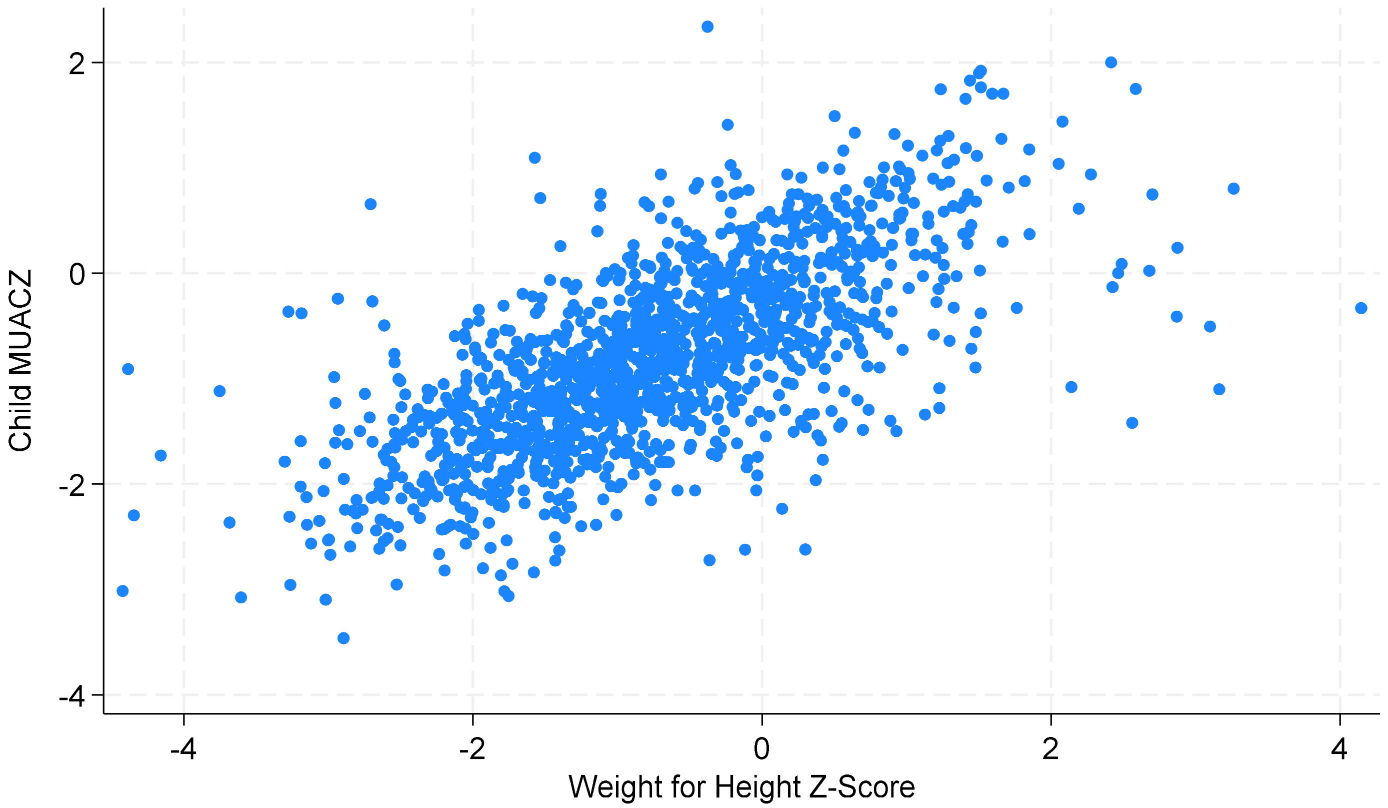

#### Annex S10. Midline Analysis Linear Regression Modelling Results

##### Table S10A. Regression Modelling for Endline Sample

| | **Model 1** β (95% CI) | **Model 2** β (95% CI) | **Model 3** β (95% CI) | **Model 4** β (95% CI) | **Model 5** β (95% CI) | **Model 6** β (95% CI) | **Model 7** β (95% CI) | **Model 8** β (95% CI) |
|---|---|---|---|---|---|---|---|---|
| MUAC | 0.59*** (0.55, 0.63) | 0.70*** (0.66, 0.74) | 0.72*** (0.68, 0.76) | 0.72*** (0.68, 0.76) | 0.70*** (0.66, 0.74) | - | - | 0.71*** (0.67, 0.75) |
| Age (continuous) | - | (-0.033)*** (-0.037, -0.030) | -0.032*** (-0.035,-0.029) | -0.030*** (-0.033, -0.026) | -0.028*** (-0.032, -0.025) | - | -0.001 (-0.004, 0.002) | - |
| Sex (Female) | - | - | 0.19*** (0.11, 0.27) | 0.33*** (0.18, 0.48) | 0.32*** (0.17, 0.47) | - | 0.072 (-0.008, 0.152) | 0.18*** (0.10, 0.26) |
| Stunting (Stunted vs. not stunted) | - | - | 0.48*** (0.37, 0.58) | 0.48*** (0.38, 0.58) | 0.43*** (0.32, 0.54) | - | 0.44*** (0.34, 0.55) | 0.49*** (0.38, 0.59) |
| Age * Sex | - | - | - | -0.18* (-0.34, -0.01) | -0.18* (-0.35, -0.02) | - | - | - |
| Region (Bay vs Hiran) | - | - | - | - | 0.15** (0.064, 0.24) | | 0.16*** (0.07, 0.24) | - |
| MUACZ | - | - | - | - | - | 0.87*** (0.82, 0.92) | 0.85*** (0.80, 0.90) | - |
| Age (reference category 9-23 months) | - | - | - | - | - | - | - | - |
| 24-35 months | - | - | - | - | - | - | - | -0.45 (-0.57, -0.33) |
| 36-47 months | - | - | - | - | - | - | - | -0.79 (-0.91, -0.66) |
| 48-59 months | - | - | - | - | - | - | - | -1.14 (-1.27, -1.71) |
| **Model R²** | **0.3156** | **0.4664** | **0.5009** | **0.5025** | **0.5063** | **0.4758** | **0.5133** | **0.4899** |

Note: MUAC and MUACZ were modelled as continuous variables. Sex, stunting, and region were modelled as dichotomous variables. Age was modelled as both a continuous variable and a categorical variable, depending on the model. *p<0.05. **p<0.01. ***p<0.001.
Note on Model 8: Linear combinations for this regression model indicated that all categories were statistically significant from each other and all p<0.001.

#### Annex S11. Midline Analysis Concordance Results

**Table S11A. Entire Sample (N=1,482) Concordance Pairs: Wasted vs. Not Wasted**

| Pair of Wasting Indicators | Kappa | Strength of Concordance |
|---|---|---|
| MUAC & WHZ | 0.0662 | None-slight |
| MUAC & MUACZ | 0.2102 | Fair |
| MUACZ & WHZ | 0.3871 | Fair |

**Table S11B. Entire Sample (N=1,482) Concordance Pairs: Normal vs. MAM vs. SAM**

| Pair of Wasting Indicators | Kappa | Strength of Concordance |
|---|---|---|
| MUAC & WHZ | 0.0487 | None-slight |
| MUAC & MUACZ | 0.1681 | Fair |
| MUACZ & WHZ | 0.3499 | Fair |

**Table S11C. Concordance Pairs Stratified by Region: Wasted vs. Not Wasted**

| Pair of Wasting Indicators | Bay Kappa | Hiran Kappa | Total |
|---|---|---|---|
| MUAC & WHZ | 0.1973 | 0.0344 | 0.0662 |
| MUAC & MUACZ | 0.2228 | 0.2048 | 0.2102 |
| MUACZ & WHZ | 0.3693 | 0.3752 | 0.3871 |

**Table S11D. Concordance Pairs Stratified by Age (9-23 months vs 24-59 months): Wasted vs. Not Wasted**

| Pair of Wasting Indicators | 9-23 months Kappa | 24-59 months Kappa | Total |
|---|---|---|---|
| MUAC & WHZ | 0.2663 | 0.0166 | 0.0662 |
| MUAC & MUACZ | 0.7197 | 0.0827 | 0.2102 |
| MUACZ & WHZ | 0.3614 | 0.3909 | 0.3871 |

**Table S11E. Number of Children Diagnosed as Wasted Using MUAC and/or WHZ Criteria**

| | | Child Wasting by WHZ | | |
|---|---|---|---|---|
| | | Not wasted | Wasted | **Total** |
| **Child Wasting by MUAC** | Not wasted | 1284 | 180 | 1464 |
| | Wasted | 9 | 9 | 18 |
| | **Total** | 1293 | 189 | 1482 |

**Table S11F. Number of Children Diagnosed as Normal vs. MAM vs. SAM Using MUAC and/or WHZ Criteria**

| | | Child Wasting by WHZ | | | |
|---|---|---|---|---|---|
| | | SAM | MAM | Normal | **Total** |
| **Child Wasting by MUAC** | SAM | 0 | 2 | 0 | 2 |
| | MAM | 2 | 5 | 9 | 16 |
| | Normal | 20 | 160 | 1284 | 1464 |
| | **Total** | 22 | 167 | 1293 | 1482 |

**Table S11G. Number of Children Diagnosed as Wasted Using MUAC and/or MUACZ Criteria**

| | | Child Wasting by MUACZ | | |
|---|---|---|---|---|
| | | Not wasted | Wasted | **Total** |
| **Child Wasting by MUAC** | Not wasted | 1357 | 107 | 1464 |
| | Wasted | 2 | 16 | 18 |
| | **Total** | 1359 | 123 | 1482 |

**Table S11H. Number of Children Diagnosed as Normal vs. MAM vs. SAM Using MUAC and/or MUACZ Criteria**

**Table S11I. Number of Children Diagnosed as Wasted Using MUACZ and/or WHZ Criteria**

| | | Child Wasting by WHZ | | |
|---|---|---|---|---|
| | | Not wasted | Wasted | **Total** |
| **Child Wasting by MUACZ** | Not wasted | 1240 | 119 | 1359 |
| | Wasted | 53 | 70 | 123 |
| | **Total** | 1293 | 189 | 1482 |

**Table S11J. Number of Children Diagnosed as Normal vs. MAM vs. SAM Using MUACZ and/or WHZ Criteria**

| | | Child Wasting by WHZ | | | |
|---|---|---|---|---|---|
| | | SAM | MAM | Normal | **Total** |
| **Child Wasting by MUACZ** | SAM | 3 | 1 | 2 | 6 |
| | MAM | 11 | 55 | 51 | 117 |
| | Normal | 8 | 111 | 1240 | 1359 |
| | **Total** | 22 | 167 | 1293 | 1482 |

#### Annex S12. Midline Full Sample ROC Analysis

##### Table S12A. Current MUAC Thresholds, Sensitivity, Specificity, AUC – Midline

| Full Sample (N=1,482) | | | |
|---|---|---|---|
| **Current Threshold** | **Sensitivity** | **Specificity** | **AUC** |
| MUAC < 12.5cm | 4.76% | 99.30% | 0.520 |
| MUACZ < -2 SD | 37.04% | 95.90% | 0.665 |

##### Table S12B. Ideal MUAC Thresholds, Sensitivity, Specificity, AUC – Midline

| Age Category | Ideal MUAC Threshold | Sensitivity | Specificity | AUC |
|---|---|---|---|---|
| Total Sample | 14.3cm | 76.19% | 69.06% | 0.726 |
| 9-23 months (n=303) | 13.7cm | 93.75% | 67.90% | 0.808 |
| 24-35 months (n=431) | 14.2cm | 79.49% | 73.98% | 0.767 |
| 36-47 months (n=374) | 14.6cm | 88.00% | 65.12% | 0.766 |
| 48-59 months (n=374) | 14.7cm | 92.65% | 65.36% | 0.790 |

##### Table S12C. Ideal MUACZ Thresholds, Sensitivity, Specificity, AUC - Midline

| Age Category | Ideal MUACZ Threshold | Sensitivity | Specificity | AUC |
|---|---|---|---|---|
| Total Sample | -1.1 SD | 88.89% | 68.21% | 0.786 |
| 9-23 months (n=303) | -1 SD | 93.75% | 74.91% | 0.843 |
| 24-35 months (n=431) | -1.1 SD | 82.05% | 73.98% | 0.780 |
| 36-47 months (n=374) | -1.2 SD | 82.00% | 70.37% | 0.762 |
| 48-59 months (n=374) | -1.3 SD | 89.71% | 65.69% | 0.777 |

#### Annex S13. Stratified Analysis: by Region - Midline

##### Table S13A. Wasting Prevalence by Region

| **Indicator** | **Bay** (n=613) | **Hiran** (n=869) | **p-values** |
|---|---|---|---|
| MUAC | **0.98%** (n=6)<br>(95% CI: 0.20%, 1.76%) | **1.38%** (n=12)<br>(95% CI: 0.61%, 2.16%) | 0.486 |
| WHZ | **5.22%** (n=32)<br>(95% CI: 3.46%, 6.98%) | **18.01%** (n=157)<br>(95% CI: 15.51%, 20.62%) | <0.001* |
| Oedema | **1.38%** (n=12)<br>(95% CI: 0.79%,2.42%) | **0.33%** (n=2)<br>(95% CI: 0.08%, 1.30%] | 0.039 |
| WHO Definition Wasted:<br>by MUAC or WHZ or Oedema | **5.87%** (n=36)<br>(95% CI: 4.01%, 7.73%) | **19.91%** (n=173)<br>(95% CI: 17.25%, 22.56%) | <0.001* |
| MUACZ | **4.57%** (n=28)<br>(95% CI: 2.91%, 6.22%) | **10.93%** (n=95)<br>(95% CI: 8.86%, 13.01%) | <0.001* |
| Wasting by all 4 Measures:<br>by MUAC, WHZ, Oedema, or MUACZ | **8.32%** (n=51)<br>(95% CI: 6.13%, 10.51%) | **23.36%** (n=203)<br>(95% CI: 20.55%, 26.17%) | <0.001* |

*p<0.05.

##### Table S13B. ROC Analysis by Region

| **ROC Analysis Results** | **Bay** | **Hiran** |
|---|---|---|
| Ideal MUAC Threshold | 14.4cm | 14.3cm |
| MUAC AUC | 0.7644 | 0.7108 |
| MUAC Sensitivity | 84.38% | 75.16% |
| MUAC Specificity | 68.50% | 66.99% |
| Ideal MUACZ Threshold | -0.9 SD | -1.2 SD |
| MUACZ AUC | 0.7939 | 0.7589 |
| MUACZ Sensitivity | 90.62% | 84.08% |
| MUACZ Specificity | 68.16% | 67.70% |

#### Annex S14. Stratified Analysis: by Child Age (9-23 mo. vs 24-59 mo.) - Midline

##### Table S14A. Wasting Prevalences by Child Age (9-23 months vs 24-59 months)

| **Indicator** | **9-23 months** (n=303) | **24-59 months** (n=1,179) | **p-values** |
|---|---|---|---|
| MUAC | **4.29%** (n=13)<br>(95% CI: 2.01%, 6.57%) | **0.42%** (n=5)<br>(95% CI: 0.05%, 0.80%) | <0.001* |
| WHZ | **10.56%** (n=32)<br>(95% CI: 7.10%, 14.02%) | **13.32%** (n=157)<br>(95% CI: 11.38%, 15.26%) | 0.200 |
| Oedema | **0.33%** (n=1)<br>(95% CI: 0.05%, 2.32%) | **1.10%** (n=13)<br>(95% CI: 0.64%, 1.89%) | 0.215 |
| WHO Definition Wasted:<br>by MUAC or WHZ or Oedema | **12.54%** (n=38)<br>(95% CI: 8.81%, 16.27%) | **14.50%** (n=171)<br>(95% CI: 12.49%, 16.51%) | 0.381 |
| MUACZ | **5.61%** (n=17)<br>(95% CI: 3.02%, 8.20%) | **8.99%** (n=106)<br>(95% CI: 7.36%, 10.62%) | 0.057 |
| Wasting by all 4 Measures:<br>by MUAC, WHZ, Oedema, or MUACZ | **13.20%** (n=40)<br>(95% CI: 9.39%, 17.01%) | **18.15%** (n=214)<br>(95% CI: 15.95%, 20.35%) | 0.041* |

*p<0.05.

##### Table S14B. ROC Analysis by Child Age (9-23 months vs 24-59 months)

| **ROC Analysis Results** | **9-23 months** | **24-59 months** |
|---|---|---|
| Ideal MUAC Threshold | 13.7cm | 14.5cm |
| MUAC AUC | 0.8082 | 0.7428 |
| MUAC Sensitivity | 93.75% | 82.80% |
| MUAC Specificity | 67.90% | 65.75% |
| Ideal MUACZ Threshold | -1 SD | -1.1 SD |
| MUACZ AUC | 0.8433 | 0.7741 |
| MUACZ Sensitivity | 93.75% | 89.71% |
| MUACZ Specificity | 74.91% | 65.66% |

***Table of Contents***

Annex S15. Description of Bay and Hiran Regions of Somalia
Annex S16. Detailed Description of CashPlus for Nutrition Study Protocol
Table S17. Study Outcome Definitions for Acute Malnutrition by Anthropometric Indicator
Annex S18. Additional Methods: Analysis Approach

**Annex S15. Description of Bay and Hiran Regions of Somalia**

The IPC estimates that 1.25 million people are living in Bay, 37% of whom are living in crisis to emergency phases of food insecurity. The estimated population of Hiran is 500,000, 20% of whom are living in crisis to emergency phases of food insecurity. According to the United Nations Refugee Agency, in 2023, there were 555,000 new displacements from Hiran primarily due to flooding, and 276,000 new displacements from Bay due to both drought and flooding. These two regions are also receiving a large proportion of displaced persons from other regions in Somalia.

**Annex S16. Detailed Description of CashPlus for Nutrition Study Protocol**

The CashPlus for Nutrition Program, funded by the Bureau for Humanitarian Assistance, is a 6-month humanitarian program in Somalia. Save the Children is implementing this program and Johns Hopkins University is studying the effectiveness and cost-effectiveness of 3 cash interventions for preventing malnutrition (wasting) among children under 5 years of age and their mothers. The study was conducted from May 2023 to January 2024 in Bay and Hiran, two regions of Somalia with high wasting prevalence. Villages were selected for the study based on size and location and participants were cluster-randomized at the village level. Eligible participants were mothers of CU5 who were enrolled in the BHA program, were not wasted by MUAC in the last year, and had no wasting by MUAC at baseline data collection in May-June 2023. Cash amounts for study arms were determined based on the Minimum Expenditure Basket, the amount needed to meet an average household’s nutritional needs for an entire month. Households in Bay received a base cash amount of $90 and households in Hiran received $70. Participants received cash monthly for 6 months. Participants were cluster-randomized into the following arms:

- Arm 1: Received Cash Assistance only
    - Bay: $90
    - Hiran: $70
- Arm 2: Received Cash + SBCC (consultations for mothers, support groups on health and nutrition topics, etc.)
    - Bay: $90
    - Hiran: $70
- Arm 3: Received Cash + Top-up Cash:
    - Bay: $90 + $35 = $125
    - Hiran: $70 + $35 = $105

Household survey data was collected from study participants at Baseline, Midline (3 months), and Endline (6 months), including questions on demographic information, household assets, food insecurity, child sickness, and diet. A sample of participants from each arm was selected to participated in qualitative focus groups to understand participants’ perspectives on program functioning and satisfaction, community attitudes towards health-seeking and wasting treatment, and how participants used their cash. Study results are currently being analyzed and prepared for publication.

**Table S17. Study Outcome Definitions for Acute Malnutrition by Anthropometric Indicator**

| Indicator | Clinical Cutoff |
|---|---|
| **MUAC** | |
| Not Wasted | MUAC ≥ 12.5 cm |
| Wasted | MUAC < 12.5 cm |
| MAM | 11.5 cm ≤ MUAC < 12.5cm |
| SAM | MUAC < 12.5cm |
| **WHZ** | |
| Not Wasted | WHZ ≥ -2 |
| Wasted | WHZ < -2 |
| MAM | -3≤ WHZ< -2 |
| SAM | WHZ < -3 |
| **MUACZ** | |
| Not Wasted | MUACZ ≥ -2 |
| Wasted | MUACZ < -2 |
| MAM | -3≤ MUACZ < -2 |
| SAM | MUACZ < -3 |
| **Oedema** | |
| Not Wasted | Absence of bilateral pitting edema |
| Wasted (SAM) | Presence of bilateral pitting edema |

MUAC: Mid Upper Arm Circumference
MAM: Moderate Acute Malnutrition
SAM: Severe Acute Malnutrition
WHZ: Weight for Height Z-score

#### Annex S18. Additional Methods: Analysis Approach

The CashPlus for Nutrition study endline data was downloaded from Kobo Toolbox (KoboCollect;v2022.4.4) and the KoboCollect application. Data was imported and cleaned in STATA. Data were recoded into numeric and string variables as needed. Categorical wasting variables were constructed using the WHO-established clinical cutoffs (found in the definitions table above). As described in the main capstone paper, data were excluded for not meeting age criteria and for biologically implausible values, leading to a final analytical dataset of 1,408 children.

We ran descriptive statistics on all demographic and anthropometric variables of interest, looking at the distribution of region, sex, age, stunting, MUAC, and z-scores in our study population. Graphs of distributions can be found in the Results Annex: all variables maintained a fairly normal distribution except for child age, with clusters around 28 and 59 months. We calculated mean, standard, deviation, median, and range for all child characteristics. To examine differences in mean characteristics by child sex, region, age categories, and wasting prevalence, we used paired t-tests and reported statistically significant results in the Results Annex. To examine differences in wasting prevalence by child characteristics like sex, age categories, and region, we used chi-squared tests: statistically significant results in the Results Annex. Wasting prevalences for each anthropometric indicator were calculated using chi-squared tests to generate percentages, 95% confidence intervals and the associated p-values.

To evaluate concordance between anthropometric indicators, we treated WHZ as an evaluative gold standard, since this criterion diagnoses the largest proportion of children as wasted and is a gold standard for wasting assessment. Tabulations of agreement between pairs of WHZ wasting, MUAC wasting, and MUACZ wasting, and their associated kappa values were reported.

ROC analyses treated WHZ as the “gold standard” and evaluated increasing thresholds of MUAC increasing by 0.1cm and MUACZ increasing by 0.1 units. These increments for MUAC were chosen to reflect the precision with which MUAC was measured in the field (to the nearest 0.1cm). MUACZ increments were chosen for ease of categorization. When evaluating what thresholds of MUAC and MUACZ were “ideal”, we first prioritized maximizing the area under the curve (AUC). When 2 thresholds produced similar AUC values, we decided to prioritize maximizing the sensitivity over specificity. Ideally we want to identify and treat as many wasted children as possible and there is no harm in providing supplemental nutrition and wasting treatment to a healthy child.
